# An immunologically active, adipose-derived extracellular matrix biomaterial for soft tissue reconstruction: concept to clinical trial

**DOI:** 10.1101/2020.10.08.20206672

**Authors:** Amy E. Anderson, Iwen Wu, Alexis J. Parrillo, David R. Maestas, Ian Graham, Ada J. Tam, Rachael M. Payne, Jeffrey Aston, Carisa M. Cooney, Patrick Byrne, Damon S. Cooney, Jennifer H. Elisseeff

**Affiliations:** Translational Tissue Engineering Center, Wilmer Eye Institute and Department of Cellular and Molecular Medicine, Johns Hopkins School of Medicine, Baltimore, MD, USA; Translational Tissue Engineering Center, Wilmer Eye Institute and Department of Biomedical Engineering, Johns Hopkins School of Medicine, Baltimore, MD, USA; Bloomberg-Kimmel Institute of Cancer Immunotherapy, Sidney Kimmel Cancer Center, Johns Hopkins School of Medicine, Baltimore, MD, USA; Department of Plastic and Reconstructive Surgery, Johns Hopkins School of Medicine, Baltimore, MD, USA; Division of Facial Plastic Surgery, Department of Otolaryngology, Johns Hopkins School of Medicine, Baltimore, MD, USA

## Abstract

Soft tissue reconstruction remains an intractable clinical challenge as current surgical options and synthetic implants may produce inadequate outcomes. Soft tissue deficits may be surgically reconstructed using autologous adipose tissue, but these procedures can lead to donor site morbidity, require multiple trips to the operating room, and have highly variable outcomes. To address the clinical need for soft tissue reconstruction, we developed an “off-the-shelf” adipose matrix from allograft human adipose tissue (acellular adipose tissue, AAT). We applied physical and chemical processing methods to remove lipids and create an injectable matrix that mimicked the properties of fat grafting materials. Biological activity was assessed using cell migration and stem cell adipogenesis assays. Characterization of the regenerative immunology properties in a murine muscle injury model revealed allograft and xenograft AAT induced pro-regenerative CD4^+^ T cells and macrophages with xenograft AAT attracting additional eosinophils secreting interleukin 4 (Il4). In immunocompromised mice, AAT injections retained similar tissue volumes as human fat grafts but did not have the cysts and calcifications that formed in the human fat graft implants. Combination of AAT with human adipose-derived stem cells (ASCs) resulted in lower implant volumes. However, tissue remodeling and new adipose development increased significantly with the addition of cells. Larger injected volumes of porcine-derived AAT demonstrated biocompatibility and greater volume retention when applied allogeneicly in Yorkshire cross pigs. Under a biologic IND application, AAT was implanted in healthy volunteers in abdominal tissue that was later removed (panniculectomy or abdominoplasty). The AAT implants were well tolerated and biocompatible in all eight human subjects. Analysis of implants removed between 1 and 18 weeks demonstrated increasing cellular infiltration and immune populations, suggesting continued tissue remodeling and the potential for long term tissue replacement.

**SUMMARY:** An adipose-derived injectable biomaterial provides volume correction for soft tissue defects while promoting pro-healing immune responses.

## INTRODUCTION

Soft tissue damage can occur due to traumatic injury, congenital and acquired medical conditions, infection, aging, or ablative surgical procedures such as tumor resection. Soft tissue defects can severely impact cosmesis and lead to functional deficits due to lack of support, reduced range of motion and scar contracture ^1^. There are limited options currently available for soft tissue repair and reconstruction. Synthetic implants can be used to treat some types of soft tissue defects such as those used for breast reconstruction following a mastectomy. However, synthetic implants do not replicate many attributes of living tissue and can induce a foreign body response. Furthermore, synthetic implants are not available for a broad range of defect sizes and anatomical locations. Surgical approaches to treat soft tissue defects may utilize autologous tissue harvested from the patient to provide a living implant for reconstruction as an alternative to synthetic implants. Adipose tissue is frequently used as a source of autologous tissue due its elasticity and availability around the body. Lipoaspirate may be collected using liposuction techniques and injected subcutaneously to fill small tissue defects, while free flap adipose tissue transfers requiring additional microvascular reconstruction are typically needed for larger volume corrections. While autologous tissue grafts provide a biological soft tissue replacement, patients experience significant variability in transplanted tissue retention in the defect site, typically losing between 40% and 60% of the original volume within 6 months. Poor viability of transplanted adipose tissue can lead to necrosis, calcifications, and cyst formation ^2-5^. The unpredictability of these procedures can result in costly secondary surgeries and are limited by the volume of autologous tissue available in each patient, as well as co-morbidities related to tissue harvest and scarring at the donor site ^6^. There is a significant need for a biomaterial solution that could provide the benefits of autologous adipose tissue with the ease of use and delivery of synthetic implants.

Biological materials can be derived from the extracellular matrix (ECM) of tissues. Tissues are processed to remove viable cells and a variety of physical, enzymatic, and chemical approaches are used to isolate and preserve the ECM in the form of sheets, particles, and gels ^7^. Biological scaffolds derived from both human allograft and swine xenograft tissue sources have been used for a variety of clinical applications since 1995 ^8^. These materials are composed of a network of structural proteins and proteoglycans that provide both mechanical support and biological cues for cell migration and tissue development. Biological cues from the ECM including growth factors and ECM-associated vesicles promote cell migration and tissue development with the implanted acellular scaffolds, gradually leading to the formation of new viable tissue with potential permanence. While autologous adipose tissue is used frequently in surgical soft tissue reconstruction, there is currently no FDA-approved biological material available that is derived from or mimics the properties of native adipose tissue.

While mobilization and activity of stem cells are considered key requirements and central mechanisms for new tissue formation in the body, recent research points to immune system engagement as a critical feature of biological scaffolds for promoting regeneration. Badylak and colleagues found that biological scaffolds derived from urinary bladder matrix tissue promoted a macrophage phenotype associated with tissue repair ^9^. This preclinical finding was supported by a clinical study of muscle repair where a decellularized biomaterial derived from small intestinal submucosa tissue resulted in a predominantly M2 (CD163^+^) macrophage phenotype within treated muscle defects ^10^. Further studies found that biological scaffolds increased recruitment of Type 2 helper T cells (T_H_2) and that these T cells were required for promoting the pro-regenerative macrophage phenotype and tissue repair ^11^. These ECM-derived biomaterials induced interleukin 4 (Il4) production, an immune-related cytokine, that is important for tissue repair in multiple tissues including muscle, liver, cartilage, and brain ^11-15^.

Here we describe the development, preclinical characterization, and pilot clinical testing of an adipose ECM biomaterial – Acellular Adipose Tissue (AAT). Previous studies characterized methods for generating AAT and evaluated rheological properties compared to commercially available fillers ^16,17^. In the present work we assessed the biological activity of the translational AAT, including its ability to promote adipose stem cell (ASC) migration and differentiation compared with a commercially available dermal tissue product. Proteomic analysis further defined differences between AAT and other ECM products. *In vivo* preclinical testing evaluated AAT biocompatibility and volume retention alone and in combination with human adipose stem cells (ASCs) with comparison to the current clinical gold standard of autologous fat grafting in athymic mice. We performed immunological characterization on both xenograft and allograft AAT in a murine model. Safety in large animals was established for an allogeneic porcine-derived AAT in swine studies, leading to the first-in-human, prospective Phase I trial.

## RESULTS

### Adipose tissue processing to create an injectable acellular extracellular matrix

To create a biological ECM scaffold material from adipose, we developed a method that included a combination of physical and chemical tissue processing amenable to production using good manufacturing practice (GMP) **(Fig. 1A**). The overall objective was to determine conditions that were mild enough to preserve the ECM as much as possible while removing cells and lipids. Further processing was performed to generate a physical structure and injection properties that were similar autologous fat grafting. Native adipose tissue (**Fig. 1B**) contains lipid-laden adipocytes (**Fig. 1C**), blood vessels, and a collagenous extracellular matrix (**Fig. 1D**). Briefly, cadaveric adipose tissue is disrupted using mechanical and chemical methods to remove intracellular lipids and cells (**Fig. 1E-F**) and is then milled into an injectable form (**Fig. 1G**). The resulting processed AAT is composed primarily of structural proteins and extracellular matrix components derived from adipose tissue along with intracellular proteins. The AAT ultrastructure resembles an interwoven, thread-like matrix of collagen and ECM proteins (**Fig. 1H-I**).

**Fig. 1.**
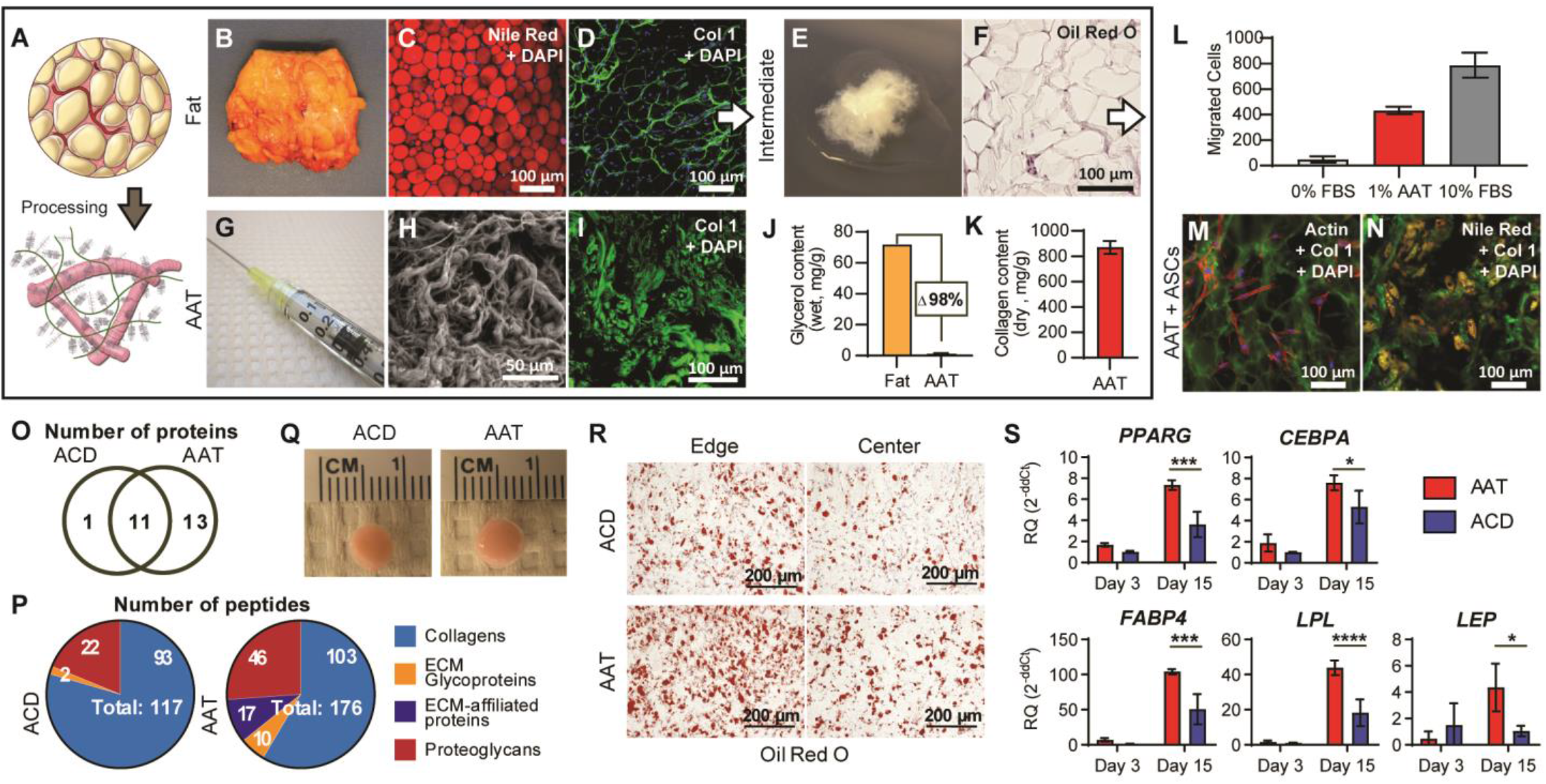
An adipose-derived biomaterial, Acellular Adipose Tissue (AAT), retains tissue-specific properties favoring *in vitro* chemo-attraction and adipogenesis. (**A**) Macroscopic representation of whole adipose tissue processing into an injectable, adipose extracellular matrix biomaterial. (**B**) Gross images of cadaveric native adipose tissue. (**C**) Nile red staining of the central lipid droplets in native adipose tissue (fat). (**D**) Fluorescent staining of the collagenous extracellular matrix (Col1) surrounding cells in native adipose tissue. (**E**) Gross image of the decellularized and delipidated processing intermediate. (**F**) Decellularized and delipidated processing intermediate stained for intracellular lipids using Oil Red O. (**G**) Gross image of the final injectable product. (**H**) Scanning electron microscopy of AAT. (**I**) Col1 fluorescent staining of AAT. (**J**) Total lipid content of AAT and matched donor adipose demonstrating >98% lipid removal in processing. (**K**) Total collagen content determined by a hydroxyproline assay. (**L**) AAT-induced migration of adipose-derived stem cells within 6 hours in a trans-well assay. (**M**) AAT reseeded with ASCs stained fluorescently for Col1 and Actin. (**N**) Nile Red staining for intracellular lipids in ASC-containing AAT supplemented with adipogenic media supplementation. (**O**) Number of shared and unique proteins between AAT and ACD by LC-MS/MS. (**P**) Categorization of peptides identified in AAT and ACD by LC-MS/MS. (**Q**) 3D constructs of ASCs cultured in ACD and AAT. (**R**) Oil Red O staining from both the edge and center of constructs and (**S**) differential gene expression for adipogenic markers within 3D constructs normalized to ACD on Day 3.

Adipocytes are large and fragile cells and their disruption can lead to leakage of intracellular lipids. These lipids have inflammatory properties that may negatively affect the host response ^4,18^ so processing conditions were optimized to eliminate as much free lipid as possible. Triglycerides account for most of the lipid in native adipose tissue, so glycerol content was used to approximate overall lipid content. Comparison of glycerol levels in AAT and matched donor adipose tissue indicate that over 98% of the intracellular lipids are removed during processing, leaving AAT with a glycerol content of less than 0.1% of its total weight (**Fig. 1J**).

Biochemical analysis determined that collagen was the primary component of AAT, 87.0% (± 5.1%) dry weight (**Fig. 1K**). To assess the composition and tissue-specific properties of AAT in more detail, we performed a liquid chromatography tandem mass spectrometry (LC-MS/MS) proteomic analysis. AAT and a commercially available dermal ECM material, Acellular Dermis (ACD, Cymetra) were digested to obtain peptides for identification by LC-MS/MS. The resulting peptide spectra were searched against a database of the human proteome to identify the proteins of origin, and were classified based on matrisome proteins as defined by Naba et al ^19^. Twenty-five matrisome or matrisome-associated proteins were identified, of which 11 proteins were common between the adipose and dermal matrices, 1 protein was unique to the dermis, and 13 were unique to adipose tissue (**Fig. 1O**).

Of the proteins shared between these two tissue products, members of the collagen family dominated the spectra with Types I, III, VI as well as proteoglycans biglycan, decorin, lumican osteoglycin and prolargin. AAT additionally contained one unique collagen (Type XIV) and two unique proteoglycans (aspirin and heparin sulfate). There was differential expression of ECM glycoproteins and ECM-affiliated proteins between the adipose and dermal ECM materials. Most ECM glycoproteins, including cartilage intermediate layer protein, dermatopontin, fibrillin, and laminin, were found only in AAT, while periostin was the only unique glycoprotein to ACD. ECM-affiliated proteins identified in AAT included annexins A1, A2, and A6 and coagulation factor XIII, whereas no ECM-affiliated proteins were detected in ACD (**Table S1**).

### AAT promotes adipose stem cell migration, adhesion, and adipogenesis *in vitro*

Tissue-derived materials provide more than a physical matrix for volume reconstruction as they contain many biological factors that can impact surrounding cells and tissues. We evaluated the chemo-attractive properties of AAT by measuring the migration of human adipose derived stem cells (ASCs) through a porous membrane in response to soluble molecular signals. Serum-starved human ASCs were screened in a Boyden chamber for 6 hours to determine whether they would migrate towards adipose ECM proteins. Serum-free media was used as the negative control for cell migration and media supplemented with 10% fetal bovine serum (FBS) was used as the positive control. A significant increase in the number of cells migrating across the membrane was observed with the addition of 1% AAT to serum-free media in the lower chamber, or 51.8% of the migration observed with the positive control of 10% FBS (**Fig. 1L**).

In addition to attracting ASCs with soluble factors, AAT provides a physical substrate for cell attachment and differentiation. ASCs seeded on AAT-coated slides preferentially adhered to the matrix proteins over areas of exposed glass and adapted a spindle-shaped mesenchymal morphology (**Fig. 1M**). With the addition of adipogenic induction media, cells underwent adipogenesis as determined by the adoption of a round morphology characteristic of mature adipocytes and accumulation of lipid droplets that stained positively with Nile Red (**Fig. 1N**).

To determine if the AAT scaffold provided a unique substrate for adipogenesis, we compared the adipogenic differentiation potential of ASCs cultured in AAT and ACD in a 3D culture environment. ASCs were suspended and cultured within AAT or ACD constructs formed in cylindrical plastic molds. Constructs were initially maintained in growth media for 3 days after which they were transferred into adipo-inductive media until day 15. After 15 days, AAT and ACD constructs looked indistinguishable upon gross examination (**Fig. 1Q**). However, histological staining with Oil Red O revealed greater lipid accumulation in the AAT construct compared to ACD (**Fig. 1R**). Furthermore, gene expression of early (PPARγ and CEPBα) and late (FABP4, LPL, LEP) markers of adipogenesis in ASCs significantly increased in the AAT compared to ACD (**Fig. 1S**).

### AAT enhances new adipose formation in combination with ASCs without viability loss observed within fat grafts

Autologous surgical methods for soft tissue reconstruction use lipoaspirate and ASCs enriched from lipoaspirate. To compare AAT to these clinical techniques, we evaluated biocompatibility and volume retention of human lipoaspirate and cells in an immune-deficient mouse model. Athymic nude mice each received subcutaneous injections of AAT with and without ASCs. For implants containing ASCs, cells were resuspended in AAT immediately prior to injection (**Fig. 2A**). No volume loss was observed in AAT without ASCs over 12 weeks (final average volume ratio of 1.08), while inclusion of ASCs in the biomaterial resulted in gradual loss of volume over the study period (final volume ratio of 0.63). Hematoxylin and eosin (H&E) staining of AAT showed extensive *de novo* adipose tissue formation and collagen remodeling where ASCs had been delivered with the biomaterial (**Fig. 2C**). In these implants, much of the AAT was replaced by new adipose tissue. The implant edges appeared completely populated by adipocytes while the center of the AAT had less adipogenesis and regions where the implant was still visible suggesting progressive inward cell migration and differentiation.

**Fig. 2.**
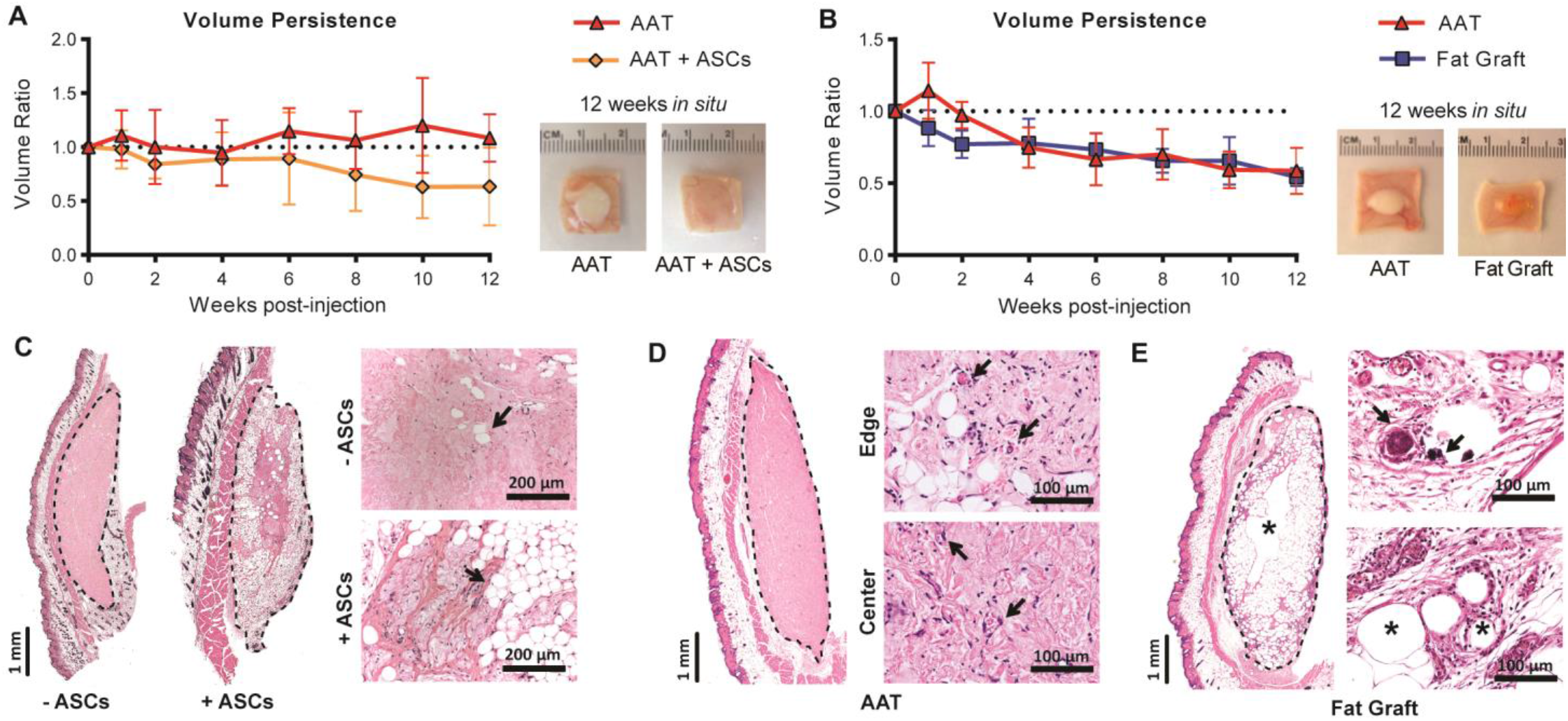
AAT demonstrates volume retention and superior biocompatibility in athymic mice. (**A**) Volume retention for human lipoaspirate (fat grafting) and AAT injected subcutaneously in athymic nude mice, as measured trans-dermally using digital calipers, with gross images of recovered implants after 12 weeks *in situ*. (**B**) Volume persistence in AAT versus in AAT supplemented with ASCs when implanted in athymic nude mice, with gross images of recovered implants after 12 weeks *in situ*. (C-E) H&E staining of implants recovered after 12 weeks, with implant boundaries indicated by dashed lines. (**C**) Adipogenesis and vascularization (arrows) within the AAT implant. (**D**) Signs of calcification (arrows) and cyst formation (*) within fat grafts. (**E**) New adipose tissue formation (arrows) within ASC-containing AAT implants.

We also compared AAT with lipoaspirate (also known as fat grafting) as the gold standard treatment for soft tissue volume correction (**Fig. 2B**). AAT and fat grafts were placed on the same mice (at distal sites) and exhibited similar loss of volume (final volume ratios of 0.59 and 0.53 for AAT and fat grafting, respectively). AAT implants lost volume faster in this study where the athymic mice also received fat grafts. AAT implants in fat-grafted animals also demonstrated increased cellular infiltration compared to AAT implanted in animals that only received the biomaterial injections (**fig. S1**).

While the AAT and fat grafts exhibited similar volume retention, gross and histological examination of the explanted AAT, AAT with ASCs, and fat grafts at 12 weeks revealed significant differences in cellular reactions at the implant site. Each type of recovered implant had a distinct gross morphology. Fat grafts retained the distinct yellow color characteristic of the intracellular lipids found in native human adipose tissue (**Fig. 2A**). AAT implants without ASCs retained the white color of the AAT material, while AAT with ASCs took on the color and texture of the surrounding tissues (**Fig. 2B**). The edge and center of AAT implants were well-infiltrated with host cells, with clusters of adipocytes present near the implant edges and small caliber blood vessels present both peripherally and in the center of implants (**Fig. 2D**). In contrast, the fat grafts had only a layer of viable adipocytes on the outside layer of the implants with large necrotic cysts towards the center of the implant. Regions of calcification surrounded by phagocytes were also observed, which typically occurs secondary to tissue necrosis (**Fig. 2E**).

### AAT induces pro-regenerative T cell and macrophage phenotypes

ECM-based biomaterials recruit immune cells and promote pro-regenerative phenotypes in macrophages and T cells. These pro-regenerative immune phenotypes are central to the biomaterial efficacy in tissue repair. Specifically, biological scaffolds induce a pro-regenerative (M2) macrophage phenotype and a CD4^+^ T helper (T_H_)2 phenotype characterized by production of Il4. Previous *in vivo* testing of AAT evaluated compatibility and efficacy of volume retention in the xenogeneic context of athymic mice and rats ^16,20^, however AAT in clinical use would constitute an allogeneic biomaterial. Since both allograft and xenograft ECM-based materials are used clinically today and both forms of AAT were assessed for volume retention and compatibility, we sought to compare their immune-modulating properties.

AAT derived from human tissue (hAAT, xenogeneic in mice) and outbred CD-1 mice (mAAT, allogeneic in mouse) were implanted in quadriceps muscle defects in Il4/GFP-enhanced transcript (4get) mice. Animals with normal (uninjured) quadriceps and injured animals treated with only saline solution served as control groups. At 7 days post-treatment, the AAT was visible in the defects and the quadriceps and associated scaffold were harvested to assess the immune responses using flow cytometry (**Fig. 3A, B**). Lymphoid and myeloid populations were quantified relative to total live cells, CD45^+^ immune cells, or total CD3^+^ T cells (**Fig. 3C**). As expected, implantation of a biomaterial significantly increased immune cell recruitment. Injury alone (saline-treated vehicle wounds) increased CD45^+^ immune cell numbers approximately 10-fold compared to healthy, uninjured muscle (40.2% vs. 3.9% of live cells). Treatment with allogeneic mAAT or xenogeneic hAAT increased immune infiltration by 1.7-fold to 2.0-fold compared to vehicle treatment alone (68.6% and 78.5% of live cells respectively). The xenogeneic hAAT recruited more than double the proportion of eosinophils (23.9% of immune cells) compared to allogeneic mAAT (10.5%), saline treatment (7.9%), or healthy muscle (8.4%). Abundance of total T cells was similar between healthy muscle and scaffold-treated injured muscles (∼5-6%), and while not statistically significant, untreated wounds had roughly half the proportion of T cells (3.2%). T cells found within both allogeneic mAAT and xenogeneic hAAT similarly skewed towards CD4^+^ T cells and away from CD8^+^ T cells relative to saline-treated wounds and healthy muscle (**Fig. 3D**). Untreated muscle wounds had a larger proportion of macrophages (40% of total immune cells vs. 6.7% in healthy muscle), while both allogeneic mAAT and hAAT contained significantly smaller proportions of macrophages compared with vehicle treatment (18.9% and 21.0% respectively).

**Fig. 3.**
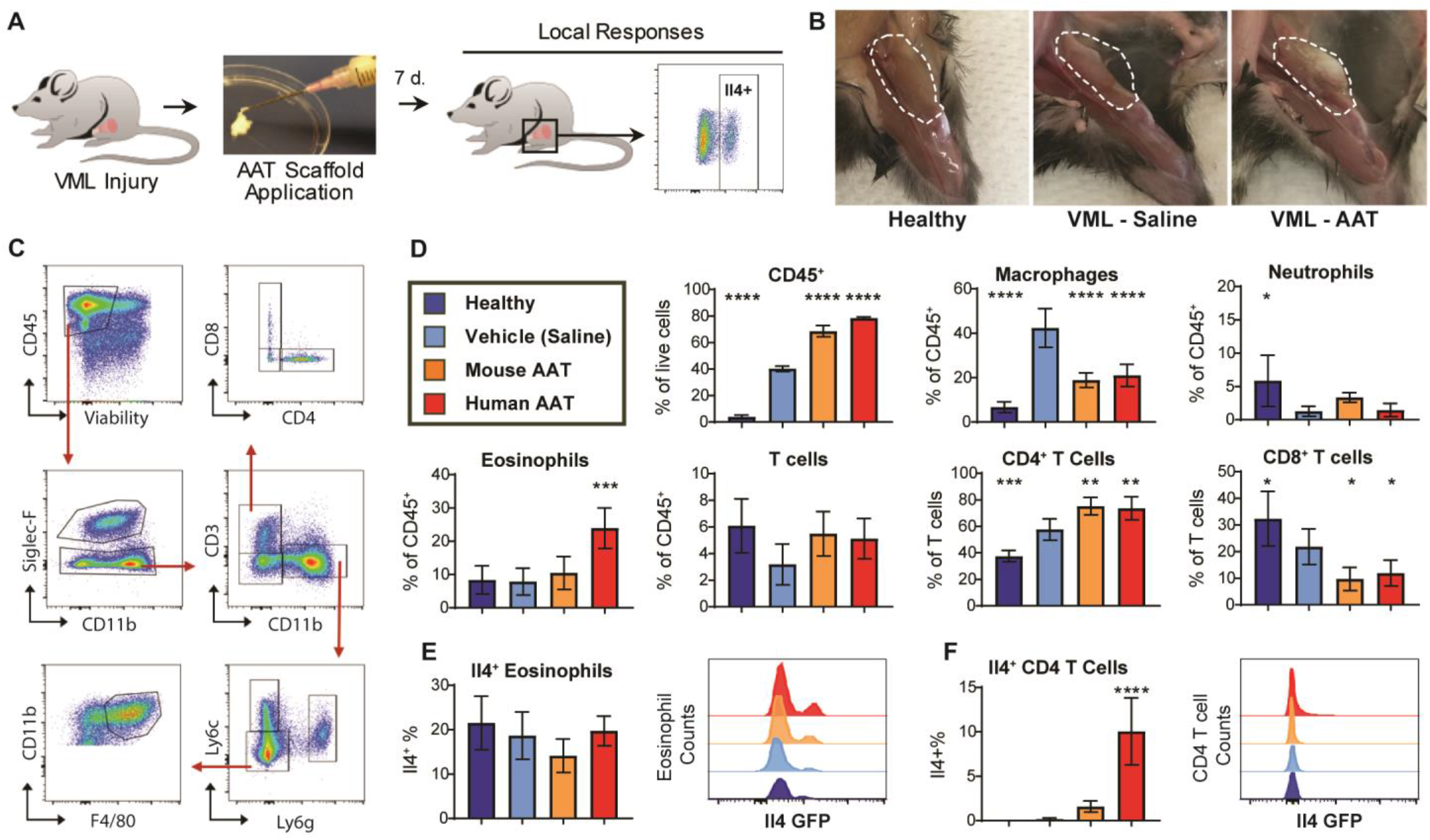
AAT elicits pro-regenerative responses in a surgical wound model of irreversible volumetric muscle loss (VML) by recruiting Il4 expressing immune cells. (**A**) Mice with bilateral surgical resections in the quadriceps muscles were treated with direct application of an AAT scaffold or saline to the wound site immediately post-op. Animals were sacrificed after 1 week to evaluate local immune responses to the biomaterials by flow cytometry. (**B**) Gross images of healthy quadriceps muscle and VML wounds treated with either vehicle (saline) or an AAT scaffold 1-week post-injury. (**C**) Flow cytometry gating of immune populations in mouse experiments. (**D**) Immune cell populations isolated from uninjured (healthy) control quadriceps muscles and muscle wounds treated with saline or an allogenic (mouse AAT, mAAT) or xenogeneic (human AAT, hAAT) ECM scaffold. (**E**) Il4 expression by percentage and median fluorescence intensity (MFI) of eosinophils and (**F**) CD4^+^ T cells isolated from wounds in 4get mice. Significance relative to vehicle control is indicated.

Il4 expression at the wound site was traced to two immune populations: eosinophils and CD4^+^ T cells. While hAAT-treated wounds contained more eosinophils than any other treatment group, the percentage of eosinophils producing Il4 was similar across all conditions (∼14-22% of total eosinophils were Il4/GFP+). Il4 GFP median fluorescence intensity (MFI) increased slightly for hAAT-derived eosinophils, but the populations are otherwise consistent in Il4 expression (**Fig. 3E**). CD4^+^ T cells recruited to scaffold-treated wounds also contributed to Il4 production at the wound, though likely to a lesser extent than eosinophils. Healthy muscle and untreated wounds contained undetectable or very low levels of Il4-expressing CD4^+^ T cells (0% and 0.16% of CD4^+^ T cells respectively). While CD4^+^ T cells in allogeneic mAAT-treated wounds had 10-fold higher Il4 expression than in saline-treated wounds (1.56% vs. 0.16% of CD4^+^ T cells), only hAAT had significant differences Il4 for this population (64.4-fold increase over saline, 10.0% of CD4^+^ T cells). MFI in the Il4^+^ CD4^+^ T cells was consistent between treatment groups (**Fig. 3F**). These findings indicate that Il4 signaling in the wound site may be enhanced by either human adipose-derived AAT specifically and/or due to its use in a xenogeneic context in this host species.

### Xenogeneic AAT induced greater eosinophil infiltration and M2 polarization compared to allogeneic AAT

We further investigated macrophage polarization with AAT treatment using the same wound model in C57BL/6 mice. We also explored whether scaffold-associated immune responses were specific to the AAT species of origin (species-matched versus species-mismatched) and evaluated an additional xenogeneic porcine adipose-derived AAT (pAAT) along with syngeneic C57BL/6 mouse adipose-derived AAT. Immune response to the syngeneic, allogeneic, and xenogeneic biomaterials in the wound site were assessed at 1- week post-treatment similarly to the 4get studies.

Scaffold-associated macrophages in the murine muscle wounds expressed markers of M1 and M2 polarization (**Fig. 4A**). Uninjured murine muscle tissue contained predominantly double positive (CD206^+^CD86^+^) macrophages but was also enriched 3.78-fold for M2 macrophages (CD206^+^CD86^-^) over M1 macrophages (CD206^-^CD86^+^). Following muscle tissue injury, macrophage phenotype shifted significantly towards a pro-inflammatory M1 polarization (M2:M1 ratio of 0.83) without affecting proportions of non-polarized or M2 polarized macrophages. All AAT materials promoted CD206 expression and suppressed CD86 expression on macrophages. This effect was more pronounced in both xenogeneic pig and human AATs (M2:M1 ratios of 14.3 and 11.9 respectively) compared to either the syngeneic or allogeneic mouse-derived AATs (M2:M1 ratios of 2.5-fold and 2.1-fold) (**Fig. 4B**). Expression of genes associated with a type 2 immune response (*Il4* and *Arg1*) increased with biomaterial treatment, though the response was significantly higher for pAAT and hAAT. Syngeneic or allogeneic AATs did not increase *IFNg* or *iNos* expression within the wounded tissues, whereas pAAT and hAAT treatments elevated expression of these type 1 genes (**Fig. 4C**).

**Fig. 4.**
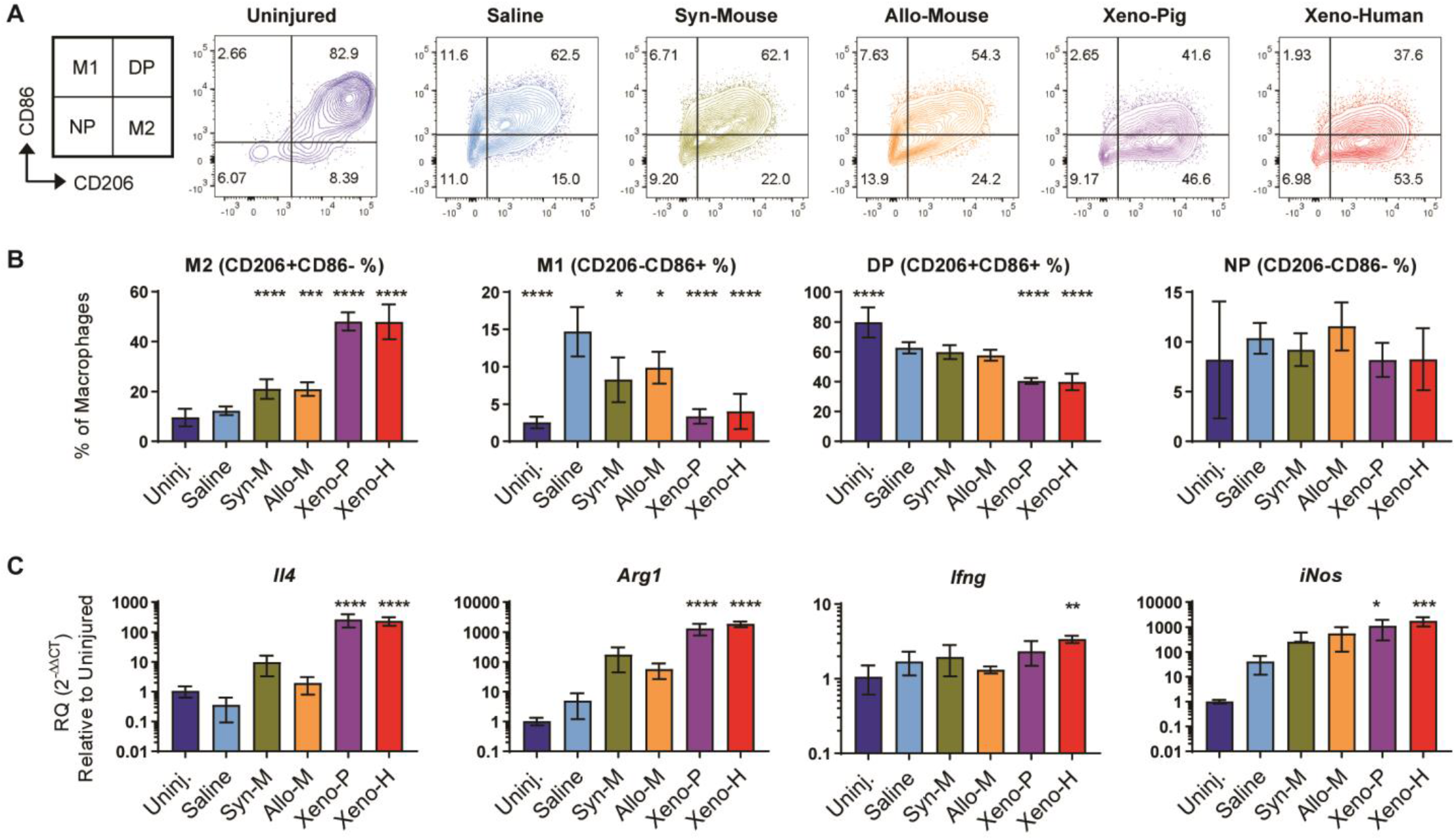
AAT scaffolds skew macrophages towards an M2 phenotype in muscle wounds. (A) CD206 and CD86 expression in macrophages isolated from healthy quadriceps muscle and scaffold- or saline-treated muscle wounds after 1-week post-op. (B) Populations of alternately activated “M2” (CD206^+^CD86^-^), classically activated “M1” (CD206^-^CD86^+^), double-polarized (CD206^+^CD86^+^) and non-polarized (CD206^-^ CD86^-^) macrophages quantified by flow cytometry. (C) Expression of canonical M2 (*Il4, Arg1*) and M1 (*Ifng, iNos*) genes at the wound site by qRT-PCR. Significance relative to vehicle control is indicated.

Overall, syngeneic and allogeneic materials behaved similarly with respect to macrophage polarization as well as recruitment of other immune cell types, and there were also no significant differences between the two xenogeneic materials in mouse models of wound healing. Both xenogeneic AATs recruited significantly more eosinophils than either syngeneic or allogeneic mAAT in mouse wounds (**fig. S2**), which likely accounts for the elevated *Il4* expression in these treatment groups and suggests that these responses are not specific to the biomaterial species of origin, but rather to how the tissue product is used. Although xenogeneic applications of AAT enhanced the magnitude of pro-regenerative immune responses, treatment with any type of AAT resulted in a significant skewing of wound-associated macrophage away from an M1- phenotype towards M2-phenotype, regardless of tissue context.

### Large injection volumes of allogeneic adipose ECM are safe and persistent in swine

Clinical soft tissue deficits can be large, requiring significant volumes to restore structure. Larger volumes of AAT were tested in Yorkshire cross pigs using cadaveric porcine adipose tissue (pAAT). Individual pigs (n = 3) each received a total volume of 48 ml of pAAT injected subcutaneously at a variety of anatomical sites with individual injection volumes up to 20 ml (**Table S2**). Implants were harvested after 4 weeks. Local responses to the allogeneic pAAT were assessed both visually and histologically. No additional swelling beyond than the original injection volume or signs of skin irritation at the injection sites were observed, and most implants were still clearly visible by external examination at the end of the study (**Fig. 5A**). A dose escalation of injection volume resulted in increased implant volumes with larger injection volumes typically translating to higher overall volume retention, although with increased variability (**Fig. 5B**). Volume retention also depended on the anatomical location of the implant (**fig. S3**). Greater resorption of implants appeared most common at sites with large amounts of native adipose or near joints where tissue compression may have occurred.

**Fig. 5.**
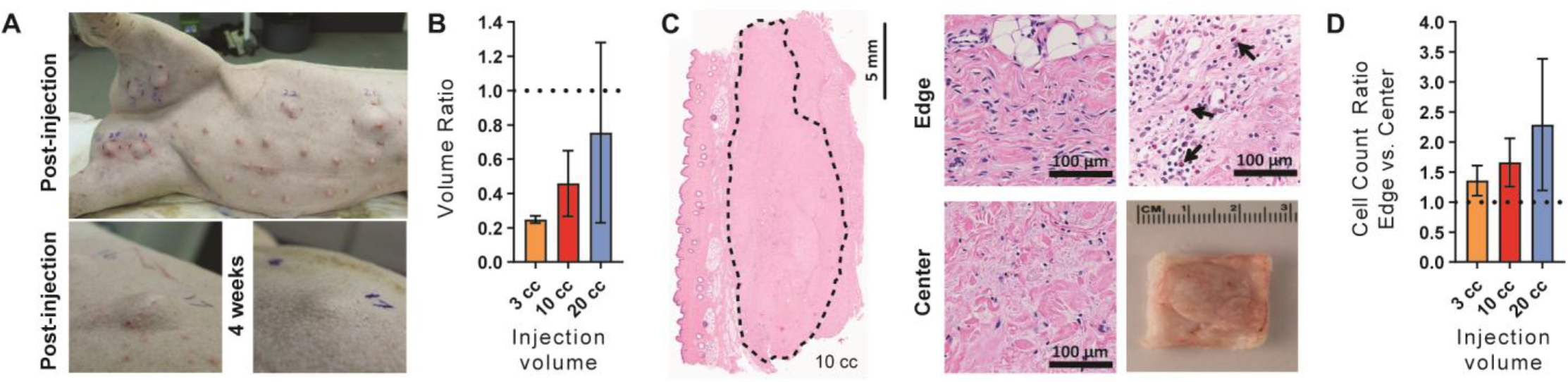
Allogenic AAT is safe and persistent at large volumes. (**A**) Porcine adipose-derived ECM implants injected subcutaneously in Yorkshire cross pigs, imaged immediately post-injection and after 1 month *in situ*. (**B**) Gross image and H&E staining of porcine-derived adipose ECM after 1 month *in situ*; high magnification images of the edge and center showing cellular infiltration, including eosinophils (arrows). (**C**) Volume retention ratio after 1 month *in situ* by initial injection volume. (**D**) Cell count ratio between edge and center of porcine implants by initial injection volume.

Histologic sections for each implant were also semi-quantitatively scored by a pathologist. Implants consisted of nonviable, acellular fibrous connective tissue matrix, few adipocytes, and a minimal to moderate immune response composed of polymorphonuclear cells (including eosinophils), lymphocytes, plasma cells, macrophages, and foreign body-type multinucleated giant cells, all predominantly at the periphery of the acellular matrix. Fibroplasia along the edge of implants was often characterized by loosely arranged fibroblasts, small caliber blood vessels, and few inflammatory cells (**Fig. 5C**). Cellular and tissue response to the implant was generally minimal to mild, and within the implant the response was generally minimal to moderate. There was some variation in the scores for implant sites from individual animals, as would be expected, but there were no substantial differences among different animals for similar implants (**Table S3- S5**). Larger implants generally contained more cells around the edges, while smaller implants had greater penetration of cells into the center of the implants (**Fig. 5D**).

### Phase I clinical study demonstrates safety and tolerability of AAT in healthy volunteers

A first-in-human, prospective Phase I trial was conducted in healthy volunteers undergoing elective tissue-removal procedures under IRB approval. The FDA designated AAT as a biologic therapy so all clinical work was performed under an Investigational New Drug application. The primary objective of this study was to assess the safety of small volume AAT injections placed in redundant tissues that would be removed surgically at increasing post-injection time points. Secondary objectives included assessments of the cellular responses within AAT implants using histological methods and the tolerability of AAT injections as reported in patient and physical satisfaction surveys. Eight subjects were enrolled in the study and received subcutaneous injection(s) of AAT in the abdomen; all implants were later excised during a panniculectomy or abdominoplasty procedure after 1 to 18-weeks *in situ* (**Table 1**).

**Table 1.** Subcutaneous injection of AAT in healthy volunteers. Participant Information

| **Table 1. Participant Information** |  |  |  |  |  |  |  |
| --- | --- | --- | --- | --- | --- | --- | --- |
| **Subject** | **Age Range** | **Sex** | **AAT Implant Time *In Situ*** | **Excision Time Point** | **Surgical Procedure** | **Injection Volumes (Sites)** | **Total Dose** |
| 01 | 60 - 69 y | M | 29 d | 4 wk | Panniculectomy | 1 cc (2) | 2 cc |
| 02* | 30 - 39 y | F | 12 d | 2 wk | Panniculectomy | 1 cc (2) | 2 cc |
| 03 | 50 - 59 y | F | 5 d | 1 wk | Abdominoplasty | 2 cc (1) | 2 cc |
| 04 | 20 - 29 y | F | 16 d | 2 wk | Abdominoplasty | 1 cc (2) | 2 cc |
| 05 | 50 - 59 y | F | 26 d | 4 wk | Panniculectomy | 1 cc (2) | 2 cc |
| 06 | 50 - 59 y | F | 6 d | 1 wk | Panniculectomy | 1 cc (2) | 2 cc |
| 07 | 40 - 49 y | F | 127 d | 18 wk | Abdominoplasty | 2 cc (1) | 2 cc |
| 08 | 60 - 69 y | F | 39 d | 6 wk | Panniculectomy | 1 cc (2), 2 cc (1) | 4 cc |
| **Mean** | **48 y** |  | **32.5 d** |  |  |  | **2.25 cc** |
| **SD** | **13.6 y** |  | **40.0 d** |  |  |  | **0.71 cc** |

\*Subject 02 withdrew from the study prior to completing the 6-week post-excision visit and 12-week post-injection PRA due to travel restrictions.

AAT demonstrated satisfactory safety results in Phase I testing with healthy volunteers (n = 8). None of the subjects experienced severe adverse events (SAEs) or unanticipated adverse events (AEs) related to the study intervention or exited the study due to an AE. Laboratory results, physical examinations, and vital signs were unremarkable throughout the study. All anticipated AEs were noted as mild and included pain/tenderness (n=2), erythema (n=4), bruising (n=4), hyperpigmentation (n=1), and textural change (n=3) at the injection site(s). Most anticipated AEs occurred during the first two weeks after injection and were resolved prior to excision (**fig. S4**). Other AEs commonly associated with injections were not observed in any participant throughout the study (no scarring, ulceration, scabbing, purpura, oozing, crusting, blanching, blistering, edema or abrasions). Volume of AAT injected did not correlate with incidence or severity of injection site reactions, suggesting that the exposures tested were within safe ranges for these individuals (**Table S6**).

Secondary outcomes of tolerability and biocompatibility were assessed globally by panel reactive antibody (PRA) testing and locally by histopathology. For 7 of 8 subjects, circulating antibodies against HLA (Human Leukocyte Antigen) proteins did not increase by either 4- or 12-weeks post-injection. One subject had a sub-clinical increase in anti-HLA antibodies at 12 weeks post-injection (11 weeks post-excision). This change is likely not related to the study intervention, as it occurred several months after removal of the implant. Therefore, we conclude none of the subjects had a systemic immunologic response which produced anti-HLA antibodies against AAT (**Table S7**).

Cellular and tissue responses near and within the implant were generally mild, typically including infiltrate of immune cells with some collagen remodeling at the implant periphery. Though accurate volume measurements could not be taken on the skin surface with calipers during this study, histological analyses reveal that significant implant volume was retained in all participants. None of the implants showed indications of encapsulation, cyst formation, or tissue necrosis (**Fig. 6A**). Overall, histological and PRA assessments revealed minimal inflammatory responses in all subjects.

**Fig. 6.**
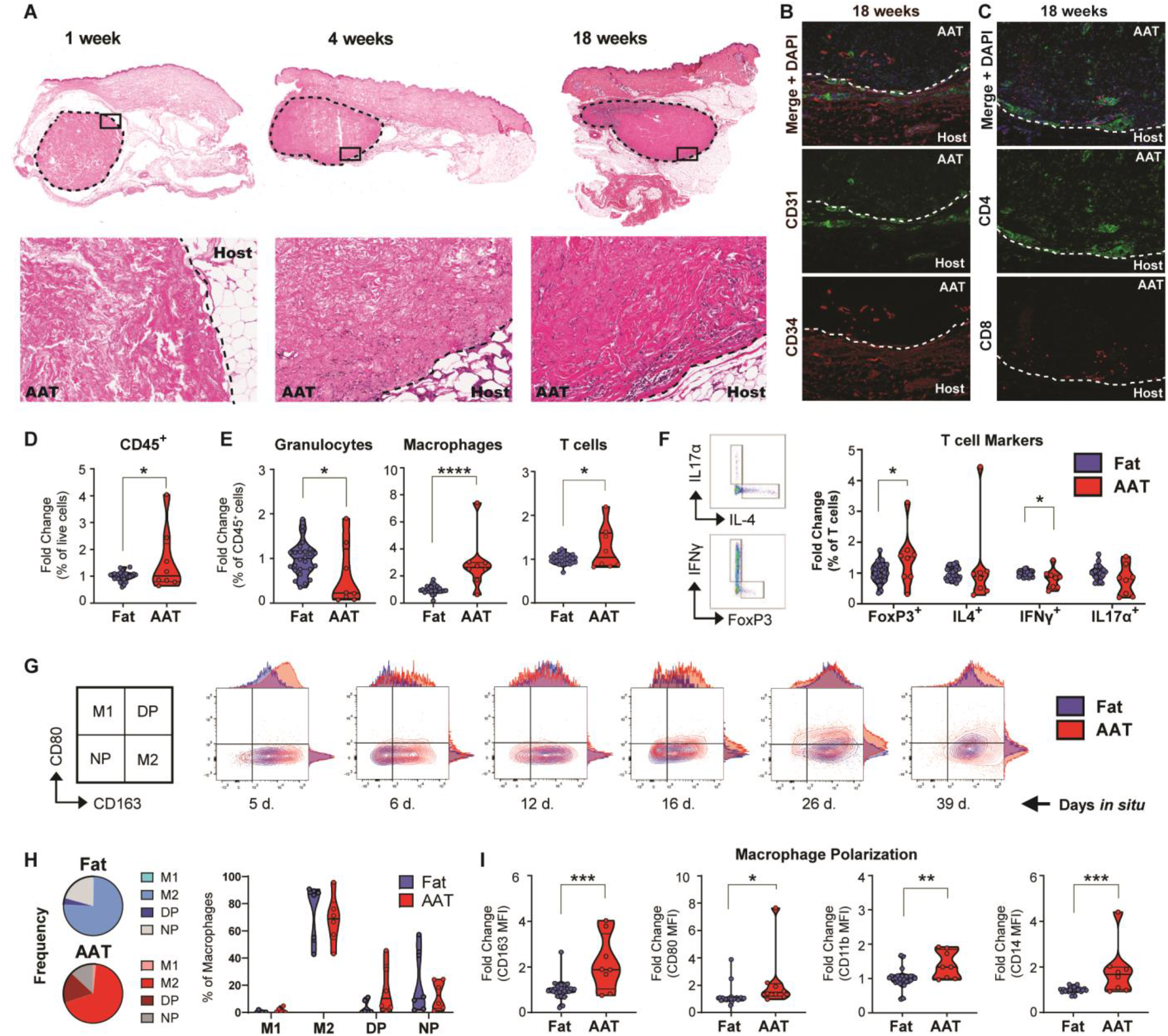
Subcutaneous injection of AAT in healthy volunteers. (**A**) H&E staining of AAT implants excised from healthy volunteers at early, mid and late time-points during a first-in-human Phase 1 study. High magnification insets show cell migration from the adjacent host tissue into the AAT matrix. (**B**) Multispectral immunohistochemistry (IHC) reveals formation of blood vessels (CD31^+^) and migration of adipose stem cells (CD34^+^). (**C**) CD4^+^ and CD8^+^ T cell infiltration by multispectral IHC. (**D**) Flow cytometry analysis was performed on recovered samples from six subjects to quantify immune cells (CD45^+^) recruited within AAT implant(s) relative to multiple samples of subject-matched adipose tissue collected distally from the injection site. Results for each recovered AAT implant were normalized to the average of multiple samples of subject-matched adipose tissue. Data shown pooled for all subjects. (**E**) Relative abundance of different immune cell subsets including granulocytes (CD11b^+^CD15^+^), macrophages (CD11b^+^CD15^-^CD11c^+^MHCII^+^CD14^+^), and T cells (CD3^+^). Data shown pooled for all subjects. (**F**) Relative expression of FoxP3 as well as T_H_1, T_H_2, and T_H_17 cytokines in T cells isolated from AAT and matched adipose tissue, shown pooled for all subjects. (**G**) Expression of macrophage polarization markers (CD163 and CD80) in AAT and matched-adipose tissue from all six individuals. (**H**) Average distribution of M1 (CD80^+^CD163^-^), M2 (CD163^+^CD80^-^), double-positive and non-polarized macrophages in AAT and matched-adipose tissue as a frequency of total macrophages. (**I**) Expression of macrophage polarization (CD163, CD80) and lineage markers (CD11b, CD14) by MFI, pooled for all subjects.

### AAT promotes local immune infiltration of CD4^+^ and FoxP3^+^ T cells and M1/M2 polarized macrophages in humans

Cellular migration from the host tissue into the implant was apparent at the host-implant boundary and increased with duration of implantation (**Fig. 6A**). After 18 weeks *in situ*, there were more cells present at the implant periphery and implant center compared to earlier time points. Multispectral immunohistochemistry also revealed the formation of new blood vessels at the edge of the implant (CD31^+^ vascular endothelial cells) and infiltration of perivascular/adipose stem cells (CD34^+^) both around and within the implant at 18 weeks post-injection (**Fig. 6B**). By this latest excision time point, CD4^+^ T cells dominated within the scaffold-recruited T cell population (**Fig. 6C**). Though both CD4^+^ and CD8^+^ T cells were found dispersed individually throughout the implant, they also often formed clusters with other non-T immune cells which resembled small tertiary lymphoid structures (**fig. S5**).

Flow cytometry was performed on tissue samples from six study participants with excision time points between 1 and 6 weeks. Immune populations present inside and around the AAT implants were quantified relative to multiple subject-matched normal adjacent adipose samples also collected at the time of implant excision (**Fig. 6D-I**). CD45^+^ immune cells (as a percentage of the total live cells) varied by subject but were generally enriched within the AAT material compared to local fat (**Fig. 6D**). Within the CD45^+^ immune compartment, granulocytes (CD45^+^CD11b^+^CD15^+^) were typically less abundant within AAT implants relative to normal fat, while macrophages and T cells were significantly enriched in AAT (**Fig. 6E**). FoxP3^+^ T cells increased within implants, representing recruitment of regulatory T cells (a subset of CD4^+^ T cells) to the AAT scaffold. Cytokine production in scaffold-associated T cells was lower than in T cells isolated from local adipose tissue, though this difference was only statistically significant across subjects for IFNγ. There was no significant difference in IL4 or IL17α production between T cells isolated from AAT versus matched adipose (**Fig. 6F**).

Polarization of scaffold-associated versus adipose-associated macrophages were assessed by CD80 and CD163 expression, markers representing classically activated (M1) and alternately activated (M2) macrophages respectively. Macrophages were identified globally by surface expression of phenotypic markers (CD45^+^CD15^-^CD11b^+^CD11c^+^CD14^+^MHCII^+^). In the six subjects tested, macrophages present in both AAT and control fat were primarily alternately activated M2 macrophages (CD163^+^CD80^-^). Recruitment within the AAT implant enhanced the M2 phenotype of scaffold-associated macrophages over fat-associated macrophages in most cases, particularly at early time points. Increases in CD80 expression were primarily found in the double-positive macrophage population (CD163^+^CD80^+^), which begins to expand at approximately 2 weeks post-injection and increases thereafter over the 6 weeks studied. AAT implants in subjects with later excision time points had larger fractions of double-positive macrophages than those with earlier excisions; similarly, early excision time points were associated with a larger proportion of unpolarized macrophages (**Fig. 6G**).

Overall, the AAT microenvironment skewed scaffold-associated macrophages towards a CD163^+^CD80^+^ double positive phenotype in AAT compared to native adipose while maintaining similar proportions of M2 and M1 macrophages (**Fig. 6H**). Scaffold associated macrophages in the AAT implants had increased expression of CD163 and CD80 (median fluorescence intensity, MFI) relative to control adipose, as well as increased expression of myeloid markers CD11b and CD14 (**Fig. 6I**).

Absolute immune cell counts within implants versus surrounding native adipose tissue were also compared for four subjects with excision time points between 1- and 6-weeks post-injection, revealing similar trends. Numbers of CD45^+^ immune cells and T cells per gram of tissue were similar between AAT and adipose tissue for all subjects. Granulocytes were consistently less abundant within AAT up to 6 weeks *in situ* than in adipose tissue, while a significant influx of macrophages occurred within the implants after 1 and 2 weeks. In these implants excised at 1 and 2 weeks (Subjects 06 and 04 respectively), the absolute number of M2 macrophages were 9.6-fold and 4.3-fold higher than in control fat. Double-polarized macrophages were even more significantly enriched within AAT at 1 and 2 weeks (565-fold and 91-fold respectively). At 4 weeks and 6 weeks, macrophage numbers were similar between AAT and local adipose tissue. Subjects also had increased absolute numbers and relative proportions of M1 (CD80^+^CD163^-^) macrophages at their injected sites; yet, M1 macrophages were by far the least abundant macrophage phenotype in all AAT implants tested (**fig. S6**).

## DISCUSSION

Adipose has been the tissue of choice for plastic and reconstructive surgeons for contouring and rebuilding soft tissue deficits. These defects can occur after oncologic resections, traumatic injuries, and as a result of congenital deformities. Autologous fat is often found in surplus and can be moved around the body in various forms such as lipoaspirate or as larger tissue grafts or adipose flap transfers. Autologous fat grafting has been used for a wide variety of clinical applications including breast reconstruction, cosmetic enhancements, and even applications in neurosurgery. Larger flap transfers (including adipose) are used for breast and extremity reconstruction and require microvascular reconstitution techniques. Because of the variability between individual patients in tissue quality, anatomical locations and sizes of adipose depots, as well as inconsistency in tissue handling or surgical methods, there can be significant uncertainty in clinical outcomes. Furthermore, the need for surgical harvesting of autologous adipose introduces donor site morbidity and challenges of inadequate tissue available for reconstruction. Insufficient donor tissue is particularly relevant, as the variability in retention often results in return trips to the operating room to harvest additional fat for grafting. Donor site morbidities may cause significant further harm to the patient ^21^ and complications include infection, necrosis, thrombosis, hematoma, and flap loss ^22,23^.

While many ECM-based products are clinically available, AAT is the first biologic therapy specifically designed to recapitulate the properties of fat grafting. A dehydrated form of adipose ECM has recently been tested in human subjects, but this product is not terminally sterilized and is not being regulated by the FDA as a biologic ^24^. Our processing methods preserve unique properties of adipose tissue including proteomic composition and rheological properties ^17^ while removing lipids that can promote inflammation. The volume maintenance of AAT appears to at least match fat grafting or stem cell delivery options, but the quality of tissue formed within AAT was significantly different than for strategies incorporating live adipocytes, particularly that calcifications and necrosis were never observed within AAT. The AAT implants in the large animal study also followed trends that are observed clinically with fat grafting ^18^. Escalating volumes of AAT were implanted safely in various anatomic locations in a swine model. Volume maintenance depended on anatomical location and larger AAT implants demonstrated greater volume retention compared to smaller implants.

Biological scaffolds derived from the extracellular matrix of different tissues are used for a variety of clinical applications. This includes products derived from dermis, small intestine submucosa, bone, and urinary bladder matrix that are approved for a variety of indications including hernia reinforcement, rotator cuff repair and wound healing. These materials are complex and defining the mechanism of action remains a challenge. The biological activity of the AAT presented here and that of other scaffolds studied is likely a combination of physical and biological cues. The proteomic analysis of AAT reveals some of its structural complexity. ECM materials also contain many cell remnants and embedded intracellular proteins that may contribute to immunological and other biological activities ^25^. Apoptotic cells are sensitive to damage signals and play an important role in immunological responses and tissue repair ^26^. Huleihel et al. discovered that extracellular vesicles containing biological factors are present within biological scaffolds and were also found in all scaffold-derived clinical products tested ^27^. Metabolic factors may also contribute to the immune phenotypes that develop in response to ECM scaffolds. Ultimately, the biological activity of ECM-based materials is well documented and likely an important factor in clinical performance. Thus, measures of activity such as cell migration or other cell function are needed to adequately predict performance of these materials.

The immune system plays a key role in the response to tissue injury and biomaterial implantation. Both innate and adaptive immune cells respond to implants and injury, and their phenotype can determine downstream repair and biocompatibility outcomes. Previous preclinical studies showed that a pro-regenerative macrophage phenotype is required for repair using biological ECM scaffolds, and that a local T_H_2 response is required for promoting the pro-regenerative macrophage phenotype that develops in response to biological scaffolds ^11^. This type 2 mediated response contrasts the macrophage and T cell phenotypes that develop around synthetic implants which includes inflammatory macrophages fusing to form foreign body giant cells and the more recently defined T_H_17 T cell and senescent cell phenotypes ^28^. The pro-regenerative or “M2” macrophage response to biological scaffolds was validated clinically in biopsies from large muscle defects treated with small intestinal submucosa-derived ECM ^29^. In the pilot clinical testing of AAT presented here, we demonstrated for the first time the infiltration of CD4^+^ T cells into the implants as observed in murine studies. At early time points, the macrophage changes associated with the pro-regenerative phenotypic response to biological scaffolds was observed. The latest time point studied, 18 weeks, demonstrated significant cellular migration evenly distributed throughout the implant.

Future studies will determine the capacity of AAT to treat larger, clinically relevant soft tissue defects. Further understanding of the immune and stromal cell changes occurring in these ECM scaffolds and related systemic changes is needed, particularly in a clinical setting that may differ significantly from preclinical models. Finally, comparison of the tissue response to xenogeneic and allogeneic ECM scaffolds in people is needed to better understand how these materials perform and to inform future technology design. While ECM scaffolds have been used clinically for years, there is still minimal understanding of their mechanisms of action and immunological effects in humans.

## MATERIALS AND METHODS

### Study Design

AAT was produced by decellularizing adipose tissue using a combination of mechanical and chemical processing steps, and the material was characterized to determine physical and biochemical properties. *In vitro* studies evaluated adipogenesis and migration of cells in response to AAT, while *in vivo* biocompatibility, volume retention, and local immune cell populations infiltrating subcutaneously injected AAT were assessed in small and large animal models. A first-in-human study determined safety and tolerability in healthy volunteers, including histocompatibility and immune-modulation at the injection site. Primary safety outcomes were evaluated in eight healthy volunteers, including adverse events (AEs), serious adverse events (SAEs) and other significant AEs, physical examination results, laboratory abnormalities, and vital signs. Biocompatibility and tolerability were assessed locally by histopathology and peripherally by panel reactive antibody (PRA) testing for circulating anti-HLA antibodies. Local infiltrating immune cells were profiling by flow cytometry in human subjects, as well as in murine studies of scaffold treated VML injuries. Materials were obtained from Sigma-Aldrich unless otherwise noted.

### Biomaterial Characterization

#### Adipose extracellular matrix preparation

For preclinical studies, subcutaneous adipose tissue was obtained from patients undergoing abdominoplasty procedures with approval from the Johns Hopkins University Institutional Review Board (IRB). Tissue was subjected to mechanical processing and extensive rinsing, followed by incubation with 3% peracetic acid for 3 hours. Samples were brought back to physiological pH using DPBS (Gibco) and incubated overnight with 1% Triton X-100 in 2 mM EDTA, followed by additional rinsing. Decellularized matrix was then snap frozen and finely cut using at Retsch GM300 knife mill. Moisture content was adjusted to 91% w/v with DPBS using an Ohas MB45 moisture analyzer. Clinical-grade AAT was manufactured from cadaveric adipose tissue under GMP conditions and terminally sterilized by gamma irradiation. Donor tissue for clinical studies was screened and acquired from a tissue bank (Donor Network West, San Ramon, CA).

#### Scanning electron microscopy

Samples were prepared for SEM as previously described ^16^ by fixing in 3.0% formaldehyde/1.5% glutaraldehyde in 0.1 M sodium cacodylate buffer with 2.5% sucrose for 1 hour. Samples were then post-fixed with 1% osmium tetroxide for 30 minutes before dehydration with graded ethanol solutions. Samples were dehydrated using CO2 critical point drying followed by sputter-coating with platinum and images were taken with FEI Quanta 200 SEM (Hillsboro, OR).

#### Lipid content

Total lipid content was determined using a triglyceride colorimetric assay. Samples were finely cut, then lipids were extracted by the Schwartz method using organic solvents and concentrated by evaporation. Enzymatic hydrolysis of triglycerides by lipase to glycerol and free fatty acids was carried out using Infinity TG Reagent (Thermo Fisher Scientific) according to the manufacturer’s protocol. Glycerol was detected by absorbance at 540nm. Absolute glycerol concentrations were determined from a standard curve and the percentage of lipids removed from AAT was determined relative to control adipose samples.

#### Collagen content

Collagen content was determined in AAT using a hydroxyproline assay kit (Sigma Aldrich) similarly to the manufacturer’s protocol. Pure collagen from bovine Achilles tendon (Sigma-Aldrich) was included as a control to calculate the number of hydroxyproline residues per molecule of collagen ^30^. AAT samples were first lyophilized, then hydrolyzed in 6 N HCl (120°C for 3 hours), diluted, transferred to assay wells, vacuum dried, then incubated at 60°C in assay reagents prior to reading absorbance at 560 nm. To eliminate any effect from endogenous interfering compounds, a collagen-spiked control sample was included to determine a correction factor. All samples were assayed at four dilutions in triplicate to calculate the mean hydroxyproline and collagen content of each sample.

#### Proteomic analysis

Adipose and dermal extracellular matrix samples were cryomilled (SPEX Sample Prep, Metuchen, NJ) and solubilized in 4 M guanidine HCl and 50 mM sodium acetate at pH 5.8, then total protein was quantified by BCA and diluted to 0.5 M guanidine HCl using distilled water. Samples were reduced in 50 mM TCEP, incubated at 37°C with vortexing for 1 hour, alkylated in 0.1 M MMTS, and incubated at room temperature for 15 minutes. Deglycosylation was carried out using deglycosylation enzyme mix (New England Biolabs) for 4 hours at 37°C. Samples were then digested overnight with 2% trypsin (Promega) at 37°C with vortexing, followed by addition of a second aliquot of 2% trypsin for an additional 4 hours.

Protein identification by liquid chromatography tandem mass spectrometry (LCMS/MS) analysis of peptides was performed using an LTQ Orbitrap Velos MS (Thermo Scientific) interfaced with a 2D nanoLC system (Eksigent). Peptides were fractionated by reverse-phase HPLC on a 75 μm x 10 cm PicoFrit column with a 15 μm emitter (PF3360-75-15-N-5, New Objective) in-house packed with Magic C18AQ (5 μm, 120Å, Michrom) using 1-45% acetonitrile/0.1% formic acid gradient over 90 min at 300 nl/min. Eluting peptides were sprayed directly into an LTQ Orbitrap Velos at 2.0 kV. Survey scans (full ms) were acquired from 350- 1800 m/z with up to 10 peptide masses (precursor ions) individually isolated with a 1.2 Da window and fragmented (MS/MS) using a collision energy of HCD35, 30s dynamic exclusion. Precursor and the fragment ions were analyzed at 30,000 and 15,000 resolutions, respectively. Peptide sequences were identified from isotopically resolved masses in MS and MS/MS spectra and searched against all human entries in RefSeq 2012, with oxidation on M, carbamidomethylation on C and Mascot Daemon (Matrix Science) software. Mass tolerances on precursor and fragment masses were 10 ppm and 0.03 Da, respectively. Mascot search result files were processed in Scaffold (Proteome Software) or Proteome Discoverer to validate protein and peptide identifications.

### In Vitro Studies

#### Adipose-derived stem cell isolation

Adipose-derived stem cells (ASCs) were isolated by digestion of fresh adipose from abdominoplasty surgical discards with 1 mg/ml collagenase I (Worthington) in DMEM F-12 (Gibco) for 1.5 hours on an orbital shaker at 37°C. The resulting cell suspension was filtered through 70 μm and 40 μm cell strainers. ASCs were seeded at 5,000 cells/cm^2^ and cultured in ASC maintenance media containing DMEM F-12, 10% fetal bovine serum (FBS, Thermo Scientific HyClone), 100 U/mL penicillin and 10 μg/mL streptomycin and passaged at 80%-90% confluency. Cells were used between passages 3-5 for all studies.

#### ASC migration assay

ASCs were serum starved for 24 hours, then trypsinized and seeded in the top chambers of a 6.5 mm / 8.0 μm pore size polystyrene transwells (Corning) at 30,000 cells per transwell. A solution of 1% (v/v) AAT in DMEM F-12 media (Life Technologies) was added in the bottom chamber. Bottom chambers containing 0% or 10% FBS in DMEM F-12 media were used as negative and positive controls. Migrated cells were quantified after 6 hours using ImageJ software (NIH).

#### Adipogenic differentiation of cells in 2D culture on AAT

AAT was embedded in OCT, cryosectioned at 200 μm and collected on a positive charge coated glass slide. OCT was removed by incubating slides in PBS on an orbital shaker with three changes each at 1-hour intervals. ASCs were seeded directly on the adipose matrix on slides or on empty glass slides and cultured in ASC maintenance media or adipogenic media. Adipogenic differentiation was carried out for ASCs seeded on the matrix with adipogenic induction media (1 μM dexamethasone, 200 μM indomethacin, 500 μM methylisobutylxanthine, 10 μg/ml insulin, 1% penicillin/streptomycin, and 10% FBS in High Glucose DMEM). Cells were differentiated for 7 days in culture before fixation for histological analysis. Slides were fixed for 10 minutes in 10% formalin and stained with Texas Red-phalloidin (Invitrogen) for actin, Nile Red for lipids, and rabbit anti-collagen type I antibodies (Fitzgerald) followed by FITC-AffiniPure goat anti-rabbit IgG secondary antibodies (Jackson) for visualization of the ECM scaffold and mounted with Vectashield plus DAPI (Vector Labs).

#### Adipo-inductive capacity in 3D culture

ASCs were resuspended in cryomilled adipose ECM or reconstituted micronized acellular dermis (Cymetra) at 2 million cells per 50 µl construct and seeded in the top chamber of a 6.5 mm transwell (Corning). After culturing in ASC maintenance media for 48 hours (10% FBS, 100 U/mL penicillin, 10 µg/mL streptomycin in DMEM F-12), constructs were transferred into adipogenic induction media (1 μM dexamethasone, 200 μM indomethacin, 500 μM methylisobutylxanthine, 10 μg/ml insulin, 1% penicillin/streptomycin, and 10% FBS in High Glucose DMEM). Cells were differentiated for 7 days in culture on an orbital shaker. For histological analysis, constructs were fixed in 10% formalin overnight, infiltrated with graded sucrose solutions, and embedded in OCT. Samples were cryosectioned at 10 μm sections and stained with hematoxylin and eosin or Oil Red O (0.75% w/v in 36% triethyl phosphate) for lipid accumulation.

#### qRT-PCR on human ASCs cultured in AAT

Snap frozen samples were homogenized in liquid nitrogen, then placed immediately placed in Trizol Reagent (Invitrogen) and total RNA extraction was carried out according to manufacturer instructions with the addition of 1 μg of glycogen added to each sample to aid in RNA precipitation. RNA quantified by Nanodrop (Thermo Scientific). cDNA was synthesized using the SuperScript RT III system (Invitrogen) and RT-PCR was carried using Power SYBR Green reagent (Applied Biosystems) on an Applied Biosystems 7500 Real-Time PCR Instrument. Each PCR reaction was carried out in triplicate with three biological replicates. Relative quantitation was performed using the ΔΔCt method ^31^ with beta actin as the housekeeping gene and normalization to expression levels in acellular dermis samples at Day 3. Primer sequences for human adipogenic genes are listed in **Table S8**.

### In Vivo Studies

All animal procedures were approved by Johns Hopkins Institutional Care and Use Committee (IACUC). All patient procedures were approved by the Johns Hopkins Institutional Review Board. Swine were housed and received veterinary care at an accredited AAALAC-1 research facility (Thomas D. Morris, Inc., Hunt Valley, MD).

#### Isolation of human lipoaspirate

Human lipoaspirate was obtained from patients undergoing liposuction. An epinephrine solution (1:500,000 in normal saline) was infiltrated into the surgical site and adipose tissue was aspirated using a 2.5 mm diameter blunt tip cannula attached to a Luer-lock syringe. Lipoaspirate was repeatedly washed with normal saline (added at a 1:1 v/v ratio) and allowed to decant at room temperature between washes to aid in removal of blood and infiltration fluids. Excess saline was removed and lipoaspirate was loaded in syringes for injection.

#### In vivo adipogenesis of AAT combined with ASCs in athymic nude mice

Female, 6-week old athymic mice (n = 12) received injections of adipose ECM with and without ASCs (2 million cells resuspended in 0.2 cc of AAT immediately prior to injection). Volume measurements were taken with digital calipers every two weeks beginning at 24 hours post-injection. Implants were removed at each of the study timepoints of 1, 4, and 12 weeks for histological analysis (n = 4).

#### Volume retention of AAT vs. lipoaspirate in athymic nude mice

Volume retention of AAT in comparison to the clinical standard of fat grafting was evaluated in 6-week old female athymic mice (n = 6). Each animal received subcutaneous injections of human lipoaspirate and AAT at discrete sites along the dorsum (0.2 cc of each material). Volume of the implanted material was measured using digital calipers immediately after injection then every two weeks until the study endpoint. After 12 weeks, implants were removed and fixed for histology.

#### Local scaffold-induced immune responses in mice with VML injuries

Mice aged 6 - 8 weeks were used to study immune responses induced by different adipose ECMs used to treat a severe muscle defect. Female wild-type C57BL/6 (Charles River) and 4get mice underwent bilateral volumetric muscle loss (VML) procedures to create a surgical defect in the quadriceps femoris muscle using previously described methods ^11^. Defects were immediately filled with 0.05 cc of either ECM material or sterile DPBS. ECM materials included AAT from xenogeneic (human or porcine), allogeneic (male outbred CD-1 mice, aged >18 weeks) and syngeneic (C57BL/6 mice) sources. After 1 week, animals were sacrificed and both quadriceps were collected for analysis.

#### Flow cytometry on scaffold-associated immune cells in mouse studies

Specimens were pooled for each individual mouse then finely diced in 1X DPBS on ice, digested for 45 minutes at 37°C in an enzyme solution consisting of 1.67 Wunsch U/ml Liberase TL (Sigma-Aldrich) and 0.2 mg/ml DNAse I (Roche) in RPMI 1640, and filtered sequentially through 100 μm and 70 µm cell strainers. Cells were stained on ice with LIVE/DEAD Fixable Aqua viability dye (Thermo Fisher) followed by a cocktail of surface markers (see Supplemental Methods). Stained cells were fixed using Cytofix reagent (BD Biosciences) and stored in DPBS supplemented with 2% FBS for up to 12 hours prior to data acquisition. Data was obtained using an LSRII flow cytometer (BD Biosciences) and analysis was conducted with FlowJo software. Gating was determined based on fluorescence minus one (FMO) plus isotype controls. Reagents were obtained from BioLegend unless otherwise noted.

#### qRT-PCR on AAT implants in murine tissue

Tissue samples collected from C57BL/6 mice were processed as described above. Relative quantitation was performed using the ΔΔCt method ^31^ with beta-2-microglobulin (B2m) as the housekeeping gene and normalization to expression levels in uninjured quadriceps muscle. Primer sequences for murine type 1 and type 2 immune response genes are listed in **Table S9**.

#### Biocompatibility and retention of large implant volumes in swine

Porcine adipose tissue (Wagner Meats, Mt. Airy MD) was processed by the same protocol as human adipose tissue to create an allogeneic AAT for swine studies. Female Yorkshire cross pigs (n = 3), aged 2.5 months, each received eight injections of allogeneic porcine-derived AAT for a total volume of 48 cc. Large single injections of 10 cc and 20 cc were placed in the flank; six smaller injections (3 cc) were placed in the ear, neck, forelimb, left and right hindlimbs, and flank. Volume measurements were taken using digital calipers immediately post-injection and at the study endpoint of 4 weeks.

#### Histology on animal tissue

Samples were retrieved from the animals (mice or pigs) and fixed in a 10% formalin solution overnight. Samples were subsequently processed with dehydration in graded ethanol solutions, cleared in xylene and paraffin embedded. Sections were cut at 5 μm and slides were stained using hematoxylin and eosin (H&E). Cell counts were quantified from a DAPI nuclear stain using ImageJ software.

### First-In-Human Testing

A Phase I clinical study was conducted at the Johns Hopkins University School of Medicine (Baltimore, MD) with approval by the Johns Hopkins University IRB.

#### Clinical trial design

An open-label pilot study was conducted in healthy volunteers undergoing elective surgery for the removal of redundant tissue. Eight participants (n = 8) were enrolled and received up to 4 cc of AAT subcutaneously in the abdomen, with 1 cc and 2 cc volumes for individual injection sites. AAT was placed during an outpatient procedure under local anesthesia using a blunt needle and following standard injection procedures. Follow-up visits (Week 1, 2, 4, and post-excision) consisted of a physician assessment of the injected area, photographic documentation of the injection sites, review of concomitant medications, and recording of any unanticipated or serious adverse events potentially associated with the injection procedure. At the end of their assigned study time point, participants had all AAT implants removed simultaneously during their elective surgery after: 1 week (±2d, n=2), 2 weeks (±2d, n=2), 4 weeks (±2d, n=2), 6 weeks (±2d, n=1) or 18 weeks (±2d, n=1) *in situ*. Abdominal tissue containing the implants was excised by the surgeon and collected by the study team. Implant(s) were excised to include a thin circumferential layer of native tissue and prepared for histopathological and flow cytometry analyses. Three to five control adipose samples (approximately 1 g each, taken >10 cm away from injection sites) were also collected from each participant.

#### Safety and tolerability

The primary outcome of safety was determined by the incidence and rate of adverse events, as well as assessment of tolerability through participant-reported comfort and physician-reported ease-of-use with the intervention. Participant blood was also screened by a panel reactive antibody (PRA) test to detect changes in levels of circulating anti-HLA (human leukocyte antigen) and other antibodies against human antigens at after treatment, which could indicate an allogeneic reaction by the subject to the AAT. PRA assessments were conducted at 4- and 12-weeks post-injection and compared to a baseline pre-injection blood draw.

#### Histopathology

Histopathological analysis of implants was performed between 1 – 18 weeks post-injection. Samples were fixed in 10% formalin, serially dehydrated in graded ethanol solutions, cleared in xylene, and embedded in paraffin. Samples were sectioned at 5 µm thickness and stained with hematoxylin and eosin (H&E). For each participant, a trained pathologist scored sections from the AAT implant and an area of distal adipose as an untreated control site.

#### Immunostaining

Multispectral immunohistochemistry was performed by sequential rounds of antigen retrieval and immunostaining using Opal Multiplex IHC reagents (Perkin Elmer). Deparaffinized and rehydrated slides were boiled in a microwave for 15 minutes in AR6 buffer (Perkin Elmer), then incubated at room temperature in 3% hydrogen peroxide for 15 minutes, blocked (4% normal goat serum/1% BSA or 10% BSA in 0.05% Tween-20 in TBS) for 30 minutes, and incubated with primary antibody for 30 minutes. Secondary detection was performed by incubating sections in MACH 4 Universal HRP-polymer (Biocare Medical) for 10 minutes, then in Opal substrate working solution for 10 minutes. Antigen retrieval and staining was then repeated for each primary antibody. Primary antibodies were purchased from Abcam and included: rabbit anti-human CD4, mouse anti-human CD8, rabbit anti-human CD31, and mouse anti-human CD34. After sequential immunostaining, sections were incubated in Opal DAPI working solution for 5 minutes and mounted in DAKO fluorescent mounting media (Agilent Technologies). Slides were stored at 4°C until imaging.

#### Flow cytometry on human tissue-derived leukocytes

Analysis of immune cell recruitment and cytokine expression was performed on dissociated AAT implants and normal adjacent tissue samples using separate panels for myeloid and lymphoid markers. Specimens were finely diced in 1X DPBS on ice, digested for 45 minutes at 37°C in an enzyme solution consisting of 1.67 Wunsch U/ml Liberase TL (Sigma-Aldrich) and 0.2 mg/ml DNAse I (Roche) in RPMI 1640, and filtered sequentially through 100 μm and 70 μm cell strainers. For intracellular staining, cells were stimulated for 4 hours at 37°C in RPMI 1640 media (Gibco) with Cell Stimulation Cocktail Plus Protein Transport Inhibitors (eBioscience) prior to staining. Cells were stained on ice with LIVE/DEAD Fixable Aqua viability dye (Thermo Fisher) followed by a cocktail of surface markers, then fixed using Cytofix reagent (BD Biosciences). For intracellular staining, cells were then permeated using Perm/Wash buffer (BD Biosciences) and stained for intracellular markers. Cells were stored in DPBS supplemented with 2% FBS for up to 12 hours prior to data acquisition. Data was obtained using an LSRII flow cytometer (BD Biosciences) and analysis was conducted with FlowJo software. Gating was determined based on fluorescence minus one (FMO) plus isotype controls. Specific antibodies included in each panel are listed in the Supplemental Methods.

#### Statistical analyses

Statistical analysis was performed using GraphPad Prism software. In grouped analyses with a single variable, significance was determined by one-way analysis of variance (ANOVA) using the Holm-Sidak correction for multiple comparisons where applicable (α = 0.05). Significance in grouped analyses with two variables was calculated using two-way ANOVA with Tukey post-hoc testing (α = 0.05). P values less than 0.05 were considered statistically significant (* < 0.05, ** < 0.01, *** < 0.001, **** < 0.0001). Plotted values represent the arithmetic mean of the data set. Error bars represent +/- one standard deviation.

## Data Availability

All data associated with this study are present in the paper or the Supplementary Materials. All primary data and analyses related to this manuscript are available upon request.

## Acknowledgements

The authors wish particularly to thank the participants in our clinical study and tissue donors who enable this research. We also thank L. Chung, K. Sadtler, and J. Andorko for flow cytometry assistance, M. Wolf for assistance with multispectral IHC assay development, C. Loveland for experimental assistance, the Sidney Kimmel Comprehensive Cancer Center (SKCCC) Flow Cytometry Core, and the Johns Hopkins Reference Histology Lab.

## Author contributions

A.E.A., I.W., and J.H.E. conceptualized these studies and drafted the manuscript. A.E.A., I.W., A.P., and J.H.E. contributed to experimental design and interpretation of results. A.E.A., I.W., and A.P. conducted experiments, analyzed data, and prepared figures. I.G. assisted with experimental procedures related to AAT development. D.R.M. performed 4get mouse breeding and assisted in experimental procedures. A.T. assisted on flow cytometry panel design and interpretation of flow data for human studies. R.P., J.A., C.C., P.B., and D.C. participated in or managed clinical activities for Phase 1 testing. All authors participated in editing and revising the manuscript text and figures.

