## Supplemental Materials for "An immunologically active, adipose-derived extracellular matrix biomaterial for soft tissue reconstruction: concept to clinical trial"

### SUPPLEMENTARY MATERIALS

#### Materials and Methods

##### Flow cytometry on dissociated murine tissues

In 4get studies, isolated cells were stained on ice with LIVE/DEAD Fixable Aqua viability dye (Thermo Fisher) followed by a surface marker cocktail: CD45 Brilliant Violet 605 (Clone 30-F11), CD11b Alexa Fluor 700 (Clone M1/70), CD3 BB700 (Clone 145-2C11, BD Biosciences), Ly-6c Brilliant Violet 510 (Clone HK1.4), Ly-6g Pacific Blue (Clone 1A8), F4/80 PE-Cy7 (Clone BM8), MHCII I-A/I-E PE-594 (Clone M5/114.15.3), Siglec-F Brilliant Violet 711 (Clone E50-2440, BD Biosciences), CD4 APC (Clone GK1.5), and CD8 PE (Clone 53-6.7). Antibodies were obtained from BioLegend unless otherwise noted.

In C57BL/6 studies, isolated cells were stained using the following cocktail of surface markers: CD45 Brilliant Violet 605 (Clone 30-F11), CD11b Alexa Fluor 700 (Clone M1/70), CD11c PerCP/Cy5.5 (Clone N418), CD3 PE-Cy5 (Clone 145-2C11), Ly-6c Brilliant Violet 510 (Clone HK1.4), Ly-6g Pacific Blue (Clone 1A8), F4/80 PE-Cy7 (Clone BM8), MHCII I-A/I-E Alexa Fluor 488 (Clone M5/114), Siglec-F PE-594 (Clone E50-2440, BD Biosciences), CD206 PE (Clone C068C2), and CD86 APC (Clone GL-1). Antibodies were obtained from BioLegend unless otherwise noted.

##### Flow cytometry on dissociated human tissues

In human flow cytometry experiments, myeloid panel surface markers included: CD45 Brilliant Violet 605 (Clone HI30), CD11b Alexa Fluor 700 (Clone M1/70), CD11c Alexa Fluor 488 (Clone 3.9), CD14 PerCP/Cy5.5 (Clone HCD14), CD15 APC (Clone W6D3), HLA-DR/DP/DQ PE-Cy7 (Clone Tu39), CD80 Brilliant Violet 421 (Clone 2D10), and CD163 PE (Clone GHI/61). Lymphoid markers were stained in a second panel of surface and intracellular markers, including: CD45 Brilliant Violet 605 (Clone HI30), CD3

Alexa Fluor 700 (Clone SK7), IL4 PE (Clone 8D4-8), IFNγ APC (Clone 4S.B3), IL17α Brilliant Violet 421 (Clone BL168), and FoxP3 Alexa Fluor 488 (Clone 150D). All antibodies were obtained from BioLegend.

### Supplemental Figures

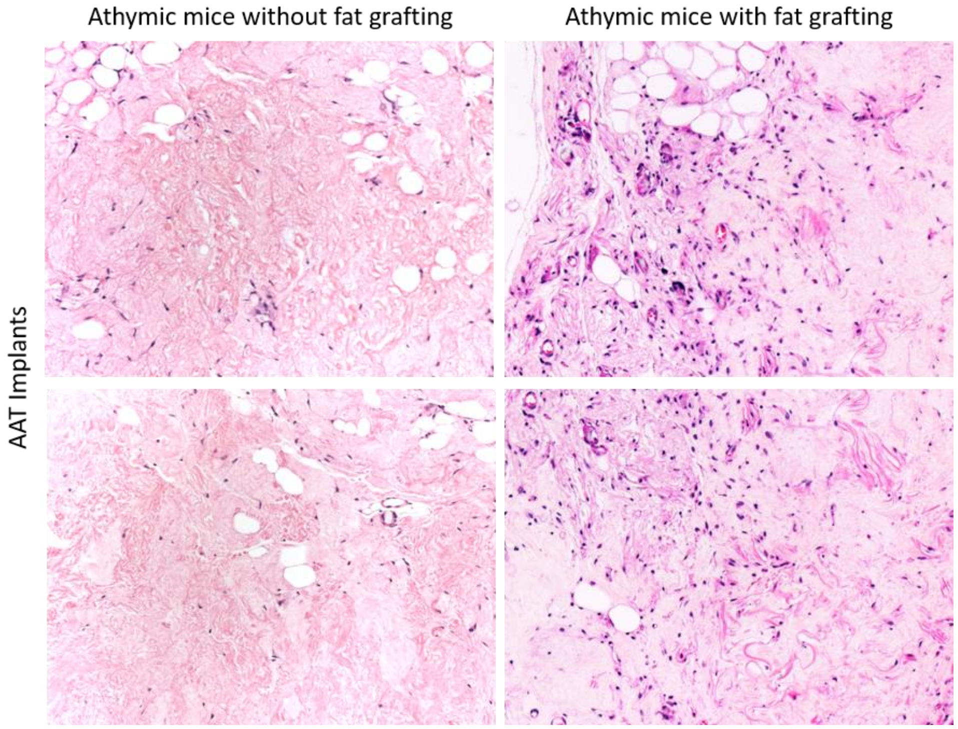


**Fig. S1**. Increased cellular infiltration of AAT implants is observed in athymic mice also injected distally with human lipoaspirate.

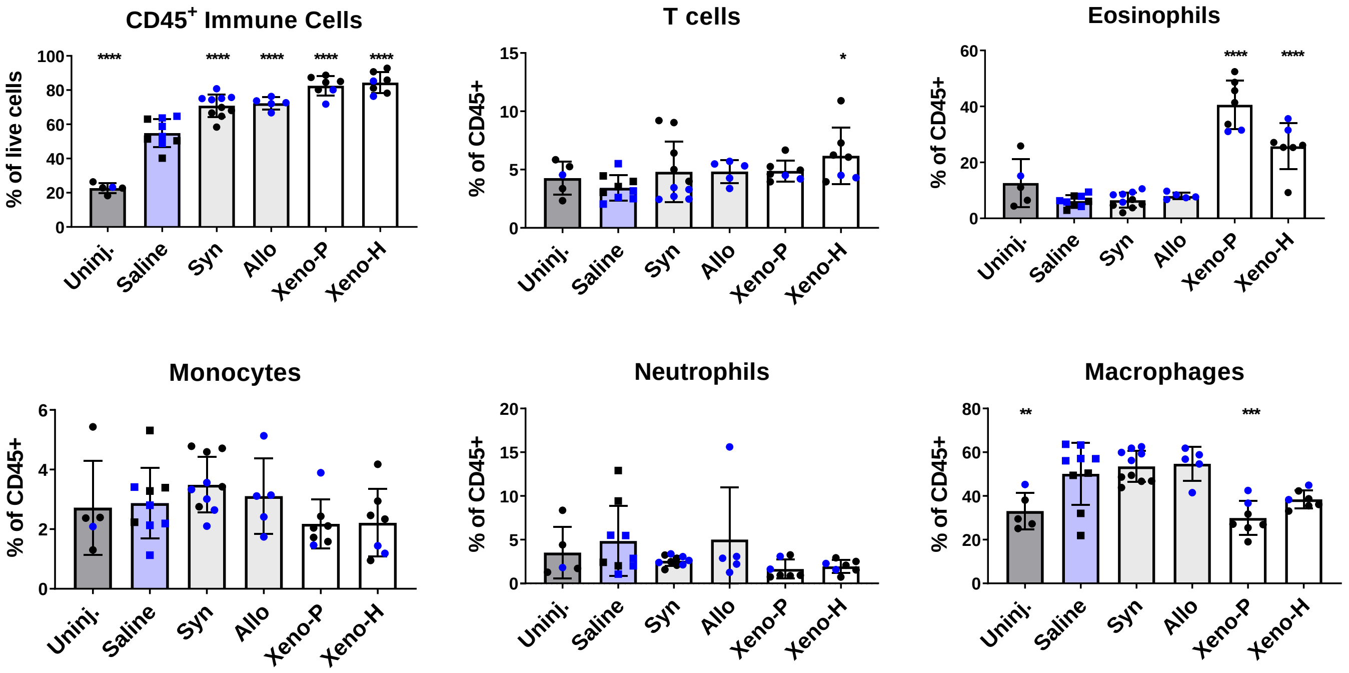


**Fig. S2.** Immune recruitment profile of syngeneic, allogeneic, and xenogeneic AATs in C57BL/6 mice with VML wounds.

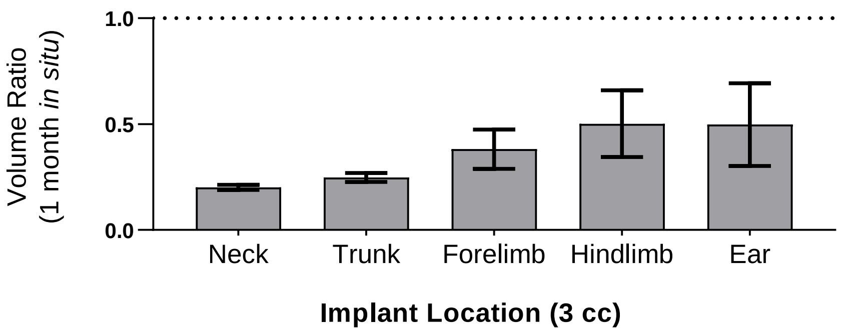


**Fig. S3**. Volume retention of pAAT implants at different anatomical sites in swine.

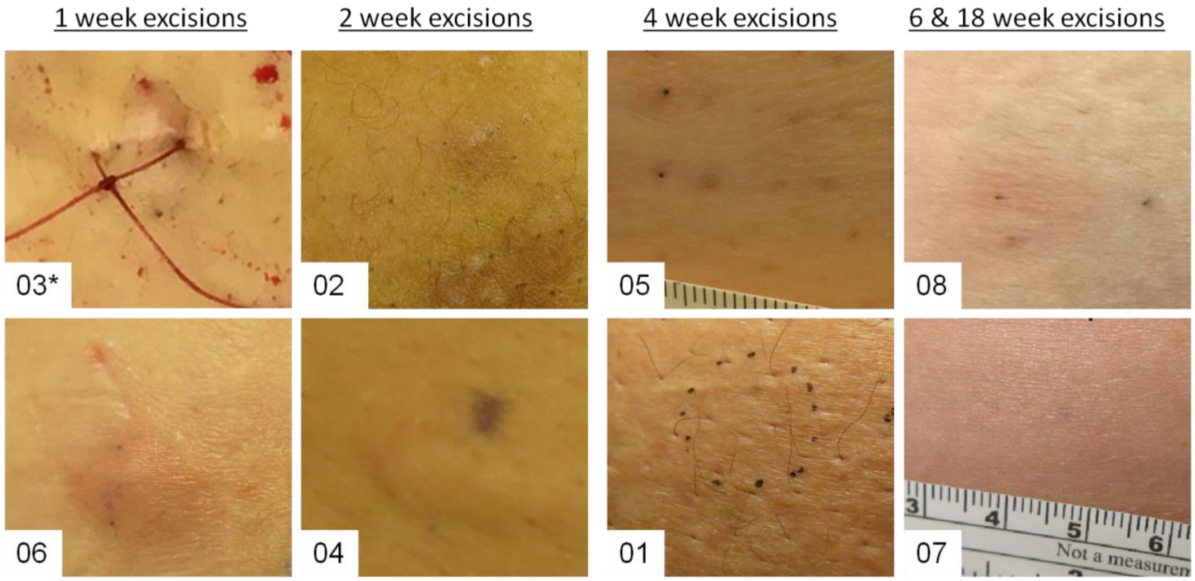


**Fig. S4.** Injection site photos at time of AAT implant excision for clinical subjects. *Excision visit photo for Subject 03 was taken after their surgery.

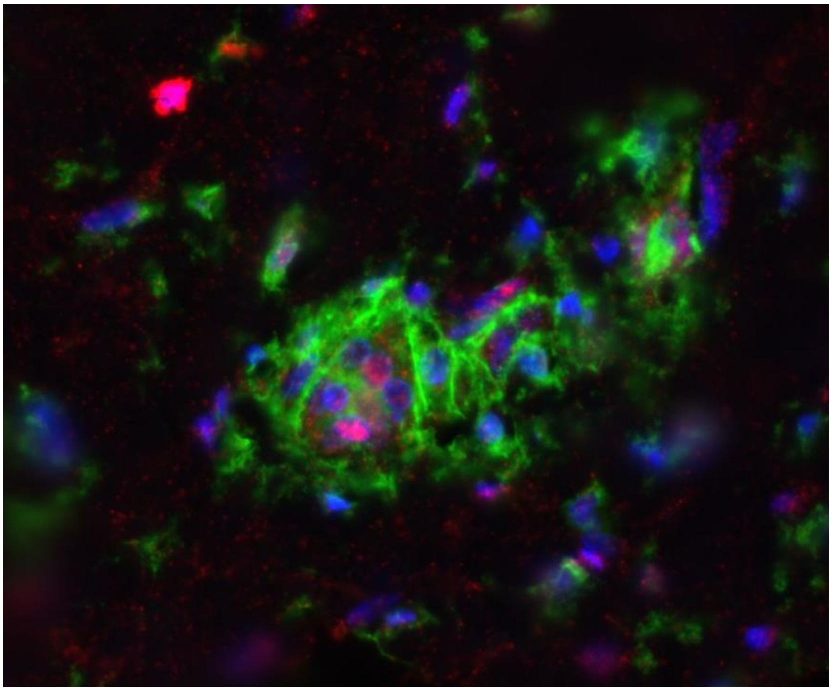

**Fig. S5**. Clusters of $CD4^+$ (green) and $CD8^+$ (red) T cells within AAT implants in human subject resemble small tertiary lymphoid structures (40x magnification, blue for DAPI).

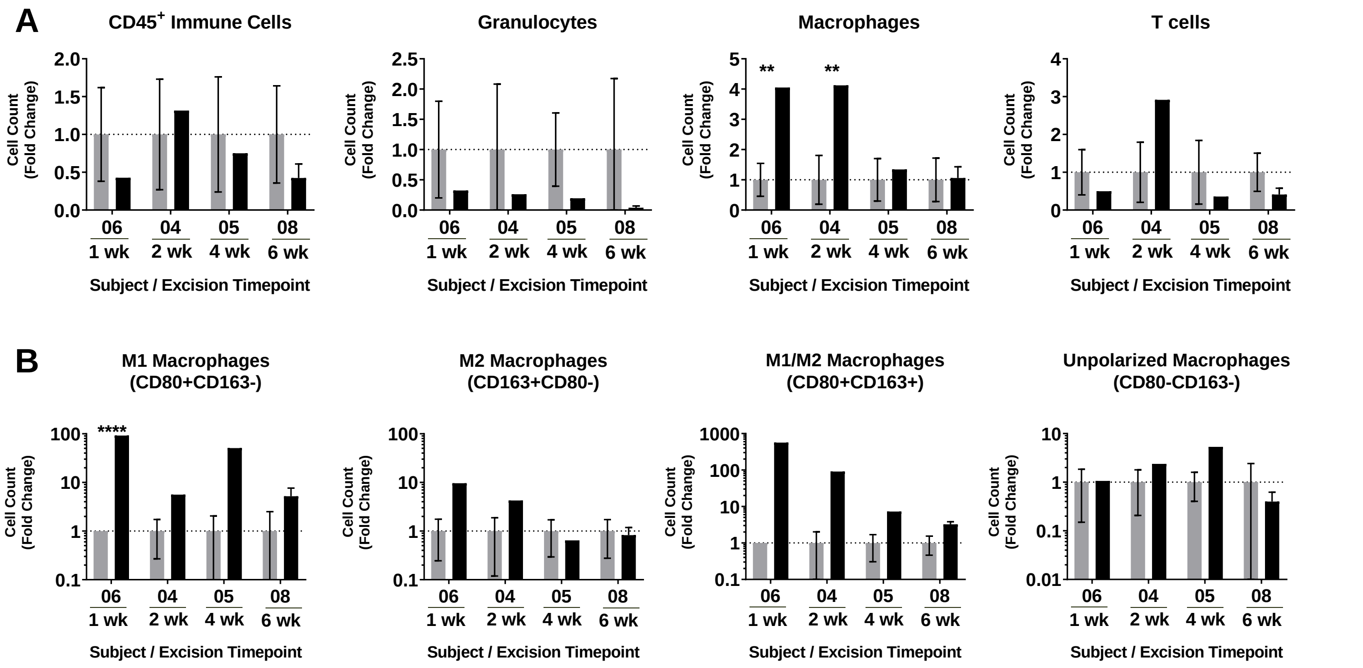


**Fig. S6**. Cell counts in human AAT implants (black bars) normalized to matched adipose tissue (grey bars) in four clinical subjects. (A) Relative quantification of total cell populations ($CD45^+$) and immune subsets, including granulocytes, macrophages, and T cells. (B) Numbers of macrophages with differential expression of polarization markers CD163 and CD80.

**Table S1**. Proteomic characterization of AAT and ACD by LC-MS/MS.

| CATEGORY | PROTEIN | Identifying peptides | | | |
|---|---|---|---|---|---|
| | | ACD 1 | ACD 2 | AAT 1 | AAT 2 |
| Proteoglycan | Asporin | 0 | 0 | 4 | 4 |
| Proteoglycan | Heparin sulfate proteoglycan 2 | 0 | 0 | 2 | 4 |
| Proteoglycan | Biglycan | 1 | 3 | 7 | 10 |
| ECM Glycoprotein | Cartilage intermediate layer protein | 0 | 0 | 1 | 2 |
| Collagen | Collagen, type I, alpha 1 | 29 | 36 | 17 | 21 |
| Collagen | Collagen, type I, alpha 2 | 21 | 16 | 8 | 9 |
| Collagen | Collagen, type III, alpha 1 | 5 | 9 | 3 | 3 |
| Collagen | Collagen, type VI, alpha 1 | 6 | 6 | 9 | 14 |
| Collagen | Collagen, type VI, alpha 2 | 3 | 8 | 7 | 9 |
| Collagen | Collagen, type VI, alpha 3 | 19 | 28 | 36 | 66 |
| Collagen | Collagen, type XIV, alpha 1 | 0 | 0 | 0 | 3 |
| Proteoglycan | Decorin | 8 | 9 | 6 | 5 |
| ECM Glycoprotein | Dermatopontin | 0 | 0 | 2 | 2 |
| ECM Glycoprotein | Fibrillin 1 | 0 | 0 | 5 | 0 |
| ECM Glycoprotein | Laminin, alpha 4 | 0 | 0 | 1 | 2 |
| ECM Glycoprotein | Laminin, beta 2 (laminin S) | 0 | 0 | 2 | 2 |
| Proteoglycan | Lumican | 0 | 2 | 5 | 12 |
| Proteoglycan | Osteoglycin | 7 | 6 | 10 | 13 |
| ECM Glycoprotein | Periostin (Osteoblast specific factor) | 1 | 2 | 0 | 0 |
| Proteoglycan | Proline/arginine-rich end leucine-rich repeat protein | 3 | 4 | 3 | 6 |
| ECM-affiliated | Annexin A1 | 0 | 0 | 4 | 4 |
| ECM-affiliated | Annexin A2 | 0 | 0 | 6 | 6 |
| ECM-affiliated | Annexin A5 | 0 | 0 | 1 | 5 |
| ECM-affiliated | Annexin A6 | 0 | 0 | 3 | 3 |
| ECM-affiliated | Coagulation factor XIII, A1 polypeptide | 0 | 0 | 0 | 2 |

**Probability Legend**

| Colour | Probability |
|---|---|
| Green | Over 95% |
| Yellow | 80% - 94% |
| Orange | 50% - 79% |

**Table S2**. Injection site locations and discrete implant volumes in safety assessment of pig-adipose derived AAT in Yorkshire cross pigs.

| Implant Number | Location | Volume (cc) |
|---|---|---|
| **1** | Right Trunk/flank | 20 |
| **2** | Left Trunk/flank | 10 |
| **3** | Left Ear | 3 |
| **4** | Left Cervical/neck, ventral | 3 |
| **5** | Left Trunk/flank | 3 |
| **6** | Left Medial forelimb | 3 |
| **7** | Left Medial hindlimb | 3 |
| **8** | Right Medial hindlimb | 3 |
| **Total Dose / Animal:** | | **48 cc** |

**Table S3**. Pathologist scoring of pig AAT implants and surrounding tissue in swine study (Pig #1).

| Study Title | Pathology Scoring: Swine Allogeneic AAT | | | | | | | |
|---|---|---|---|---|---|---|---|---|
| Time Point | 1 Month | | | | | | | |
| Animal Number | 1 | | | | | | | |
| Implant Number | 1 | 2 | 3 | 4 | 5 | 6 | 7 | 8 |
| Implant Material | Pig AAT | Pig AAT | Pig AAT | Pig AAT | Pig AAT | Pig AAT | Pig AAT | Pig AAT |
| Location | L Ear | L Cervical / neck, ventral | L Trunk/ flank | L Trunk/ flank | L Medial forelimb | L Medial hindlimb | R Trunk/ flank | R. Medial hindlimb |
| **TISSUE** | | | | | | | | |
| Cell Type/Response | | | | | | | | |
| Polymorphonuclear cells | 0 | 0 | 0 | 0 | 0 | 0 | 0 | 0 |
| Lymphocytes | 1 | 1 | 1 | 2 | 1 | 1 | 2 | 1 |
| Plasma Cells | 0 | 0 | 0 | 0 | 0 | 0 | 0 | 0 |
| Macrophages | 0 | 1 | 1 | 1 | 1 | 1 | 1 | 1 |
| Giant Cells | 0 | 0 | 0 | 0 | 0 | 0 | 0 | 0 |
| Edema Fluid Accumulation | 0 | 0 | 0 | 0 | 0 | 0 | 0 | 0 |
| Necrosis | 0 | 0 | 1M | 1M | 0 | 0 | 2M | 0 |
| Response | | | | | | | | |
| Congestion | 0 | 0 | 0 | 0 | 0 | 0 | 0 | 0 |
| Mineralization | 0 | 0 | 0 | 0 | 0 | 0 | 1M | 0 |
| Fibroplasia | 0 | 0 | 0 | 0 | 0 | 0 | 0 | 0 |
| Fibrosis | 0 | 0 | 0 | 0 | 0 | 0 | 1M | 0 |
| **IMPLANT** | | | | | | | | |
| Cell Type/Response | | | | | | | | |
| Polymorphonuclear cells | 0 | 0 | 0 | 0 | 0 | 0 | 0 | 0 |
| Lymphocytes | 2 | 2 | 2 | 2 | 3 | 2 | 3 | 3 |
| Plasma Cells | 1 | 1 | 1 | 1 | 1 | 1 | 1 | 1 |
| Macrophages | 1 | 1 | 1 | 2 | 2 | 1 | 1 | 1 |
| Giant Cells | 1 | 1 | 1 | 1 | 1 | 1 | 1 | 1 |
| Edema Fluid Accumulation | 0 | 0 | 0 | 0 | 0 | 0 | 0 | 0 |
| Nonviable acellular matrix | 0 | 0 | 0 | 2 | 1 | 0 | 3 | 0 |
| Response | | | | | | | | |
| Congestion | 0 | 0 | 0 | 0 | 0 | 0 | 0 | 0 |
| Mineralization | 0 | 0 | 0 | 0 | 0 | 0 | 0 | 0 |
| Fibroplasia | 0 | 0 | 0 | 2 | 0 | 0 | 2 | 0 |
| Fibrosis | 4 | 4 | 4 | 4 | 4 | 4 | 3 | 4 |
| Area(s) of dropout | P | NP | NP | P | P | NP | P | NP |

**Key:** P = Present, NP = Not Present, 0 = None, 1 = Minimal, 2 = Mild, 3 = Moderate, 4 = Severe, R = Rare, M = associated with skeletal muscle

**Table S4**. Pathologist scoring of pig AAT implants and surrounding tissue in swine study (Pig #2).

| Study Title | Pathology Scoring: Swine Allogeneic AAT | | | | | | | |
|---|---|---|---|---|---|---|---|---|
| Time Point | 1 Month | | | | | | | |
| Animal Number | 2 | | | | | | | |
| Implant Number | 1 | 2 | 3 | 4 | 5 | 6 | 7 | 8 |
| Implant Material | Pig AAT | Pig AAT | Pig AAT | Pig AAT | Pig AAT | Pig AAT | Pig AAT | Pig AAT |
| Location | L Ear | L Cervical / neck, ventral | L Trunk/ flank | L Trunk/ flank | L Medial forelimb | L Medial hindlimb | R Trunk/ flank | R. Medial hindlimb |
| **TISSUE** | | | | | | | | |
| Cell Type/Response | | | | | | | | |
| Polymorphonuclear cells | 0 | 0 | 0 | 0 | 0 | 0 | TM | 0 |
| Lymphocytes | 2 | 2 | 2 | 1 | 2 | 2 | TM | 1 |
| Plasma Cells | 0 | 0 | 1 | 0 | 0 | 0 | TM | 0 |
| Macrophages | 1 | 1 | 1 | 1 | 1 | 1 | TM | 1 |
| Giant Cells | 1 | 0 | 0 | 0 | 0 | 0 | TM | 0 |
| Edema Fluid Accumulation | 0 | 0 | 0 | 0 | 0 | 0 | TM | 0 |
| Necrosis | R-M | 0 | R-M | 0 | 0 | 0 | TM | 0 |
| Response | | | | | | | | |
| Congestion | 0 | 0 | 0 | 0 | 0 | 0 | TM | 0 |
| Mineralization | 0 | 0 | 0 | 0 | 0 | 0 | TM | 0 |
| Fibroplasia | 0 | 0 | 0 | 0 | 0 | 0 | TM | 0 |
| Fibrosis | 1 | 0 | 1 | 0 | 0 | 0 | TM | 0 |
| **IMPLANT** | | | | | | | | |
| Cell Type/Response | | | | | | | | |
| Polymorphonuclear cells | 0 | 0 | 0 | 0 | 0 | 0 | TM | 0 |
| Lymphocytes | 2 | 2 | 2 | 3 | 2 | 2 | TM | 2 |
| Plasma Cells | 1 | 1 | 1 | 1 | 1 | 1 | TM | 1 |
| Macrophages | 1 | 1 | 1 | 1 | 1 | 1 | TM | 1 |
| Giant Cells | 1 | 1 | 1 | 1 | 1 | 1 | TM | 1 |
| Edema Fluid Accumulation | 0 | 0 | 0 | 0 | 0 | 0 | TM | 0 |
| Nonviable acellular matrix | 1 | 0 | 0 | 2 | 0 | 1 | TM | 0 |
| Response | | | | | | | | |
| Congestion | 0 | 0 | 0 | 0 | 0 | 0 | TM | 0 |
| Mineralization | 0 | 0 | 0 | 0 | 0 | 0 | TM | 0 |
| Fibroplasia | 0 | 0 | 0 | 0 | 0 | 0 | TM | 0 |
| Fibrosis | 4 | 4 | 4 | 4 | 4 | 4 | TM | 4 |
| Area(s) of dropout | P | NP | NP | P | NP | P | TM | NP |

**Key:** P = Present, NP = Not Present, 0 = None, 1 = Minimal, 2 = Mild, 3 = Moderate, 4 = Severe, R = Rare, M = associated with skeletal muscle

**Table S5**. Pathologist scoring of pig AAT implants and surrounding tissue in swine study (Pig #3).

| Study Title | Pathology Scoring: Swine Allogeneic AAT | | | | | | | |
|---|---|---|---|---|---|---|---|---|
| Time Point | 1 Month | | | | | | | |
| Animal Number | 3 | | | | | | | |
| Implant Number | 1 | 2 | 3 | 4 | 5 | 6 | 7 | 8 |
| Implant Material | Pig AAT | Pig AAT | Pig AAT | Pig AAT | Pig AAT | Pig AAT | Pig AAT | Pig AAT |
| Location | L Ear | L Cervical / neck, ventral | L Trunk/ flank | L Trunk/ flank | L Medial forelimb | L Medial hindlimb | R Trunk/ flank | R. Medial hindlimb |
| **TISSUE** | | | | | | | | |
| Cell Type/Response | | | | | | | | |
| Polymorphonuclear cells | 0 | 0 | 0 | 0 | 0 | 0 | 1 | 0 |
| Lymphocytes | 1 | 1 | 1 | 2 | 1 | 1 | 1 | 1 |
| Plasma Cells | 0 | 0 | 0 | 1 | 0 | 0 | 0 | 0 |
| Macrophages | 1 | 1 | 1 | 1 | 1 | 1 | 1 | 1 |
| Giant Cells | 0 | 0 | 0 | R | 0 | 0 | 0 | 0 |
| Edema Fluid Accumulation | 0 | 0 | 0 | 0 | 0 | 0 | 0 | 0 |
| Necrosis | 0 | 0 | 1M | 1M | 0 | 0 | 0 | 0 |
| Response | | | | | | | | |
| Congestion | 0 | 0 | 0 | 0 | 0 | 0 | 0 | 0 |
| Mineralization | 0 | 0 | 0 | 0 | 0 | 0 | 0 | 0 |
| Fibroplasia | 0 | 0 | 0 | 0 | 0 | 0 | 0 | 0 |
| Fibrosis | 0 | 0 | 1 | 1 | 0 | 0 | 1M | 0 |
| **IMPLANT** | | | | | | | | |
| Cell Type/Response | | | | | | | | |
| Polymorphonuclear cells | 0 | 0 | 0 | 0 | 1 | 1 | 1 | 0 |
| Lymphocytes | 1 | 2 | 1 | 2 | 3 | 2 | 2 | 2 |
| Plasma Cells | 0 | 1 | 0 | 1 | 1 | 1 | 1 | 1 |
| Macrophages | 1 | 2 | 1 | 1 | 2 | 1 | 2 | 1 |
| Giant Cells | 1 | 1 | 1 | 1 | 1 | 1 | 1 | 1 |
| Edema Fluid Accumulation | 0 | 0 | 0 | 0 | 0 | 0 | 0 | 0 |
| Nonviable acellular matrix | 1 | 0 | 0 | 2 | 1 | 1 | 3 | 0 |
| Response | | | | | | | | |
| Congestion | 0 | 0 | 0 | 0 | 0 | 0 | 0 | 0 |
| Mineralization | 0 | 0 | 0 | 0 | 0 | 0 | 0 | 0 |
| Fibroplasia | 0 | 0 | 0 | 0 | 0 | 0 | 4 | 0 |
| Fibrosis | 4 | 4 | 4 | 4 | 4 | 4 | 4 | 4 |
| Area(s) of dropout | P | NP | NP | NP | P | P | P | NP |

**Key:** P = Present, NP = Not Present, 0 = None, 1 = Minimal, 2 = Mild, 3 = Moderate, 4 = Severe, R = Rare, M = associated with skeletal muscle

**Table S6**. Summary of anticipated AEs noted by study visit in Phase 1 clinical testing.

| **Excision Time Point**<br>**Subject ID** | **1 week**<br>03 | <br>06 | **2 weeks**<br>02 | <br>04 | **4 weeks**<br>01 | <br>05 | **6 wk**<br>08 | **18 wk**<br>07 | **Total** |
|---|---|---|---|---|---|---|---|---|---|
| Pain/Tenderness | | | | | Inj | | 1wk | 1wk | **3/8 (37.5%)** |
| Erythema | | Ex | Ex | 1wk | | | 1wk<br>Ex | | **4/8 (50%)** |
| Bruising | | Ex | | | | 1wk | 2wk | 1wk | **4/8 (50%)** |
| Hyperpigmentation | | | | | | | Ex | | **1/8 (12.5%)** |
| Textural change* | | | | | | 1wk<br>2wk | 1wk<br>4wk<br>Ex | 1wk<br>2wk<br>4wk | **3/8 (37.5%)** |
| Other (define) | Ex<br>Implant indurated | | | Ex<br>Implants indurated | | Ex<br>Implant raised | | | **3/8 (37.5%)** |

*Textural change was described as implant being "raised", "indurated", or "palpable"

**Study Visit Key: Inj= Injection visit, 1wk= 1-week post-injection visit, 2wk= 2-week post-injection visit, 4wk= 4-week post-injection visit, Ex= Excision visit

**Table S7**. Panel reactive antibody screening for Phase 1 subjects.

| Subject ID | Excision Time point | BASELINE TESTING<br>IgG HLA Positive | POST-INJECTION TESTING<br>Increase in IgG HLA antibodies at:<br>*4 weeks* | *12 weeks* |
|---|---|---|---|---|
| **03** | **1 week** | Yes | No | No |
| **06** | **1 week** | No | No | **Yes; lowest antibody detection level CPRA $_{Low}$ = 44** |
| **02** | **2 weeks** | **Participant withdrew prior to follow-up** | | |
| **04** | **2 weeks** | Yes | No | No |
| **05** | **4 weeks** | No | No | No |
| **01** | **4 weeks** | No | No | No |
| **08** | **6 weeks** | No | No | No |
| **07** | **18 weeks** | Yes | No | No |

**Table S8. Human qRT-PCR Primers**

| Gene | Forward Sequence 5’ – 3’ | Reverse Sequence 5’ – 3’ |
|---|---|---|
| *PPARG* | AGGAGAAGCTGTTGGCGGAGA | TGCTTTGGTCAGCGGGAAGG |
| *LPL* | GTCAGAGCCAAAAGAAGCAGCAA | GGGTTTCACTCTCAGTCCCAGAA |
| *LEP* | TGACACCAAAACCCTCATCAAGACAA | GGAGCCCAGGAATGAAGTCCAA |
| *CEBPA* | TCACCGCTCCAATGCCTACTG | CCTGCTCCCCTCCTTCTCTCAT |
| *FABP4* | ACAGGAAAGTCAAGAGCACCATAACC | TGACGCATTCCACCACCAGTTT |
| *B-ACTIN* | GGCACCCAGCACAATGAA | GCTAACAGTCCGCCTAGAAGC |

**Table S9. Murine qRT-PCR Primers**

| Gene | Forward Sequence 5’ – 3’ | Reverse Sequence 5’ – 3’ |
|---|---|---|
| *Il4* | GGTCACAGGAGAAGGGACGC | AGCACCTTGGAAGCCCTACA |
| *Ifng* | GTCAGAGCCAAAAGAAGCAGCAA | TGTCACCATCCTTTTGCCAGT |
| *Arg1* | ACAAGACAGGGCTCCTTTCAG | TAAAGCCACTGCCGTGTTCA |
| *iNos* | CTTGGTGAAGGGACTGAGCTG | GTTCTCCGTTCTCTTGCAGTTG |
| *B2m* | CACTGAATTCACCCCCACTGA | TCTCGATCCCAGTAGACGGT |
